# Personalise Dose Regimen of Vitamin D_3_ Using Physiologically-Based Pharmacokinetic Modelling

**DOI:** 10.1101/2020.12.06.20244897

**Authors:** Zhonghui Huang, Tao You

## Abstract

**Background and Aim:** Vitamin D_3_ (i.e. cholecalciferol) produces an active metabolite 25-hydroxyvitamin D_3_ (i.e. 25(OH)D_3_) to promote intestinal calcium absorption. Given high population heterogeneity in 25(OH)D_3_ plasma concentration profiles, vitamin D_3_ dose regimen needs to be personalised. The objective of this study is to establish a model that accurately predicts 25(OH)D_3_ pharmacokinetics (PK) on an individual level to enable selection of an appropriate dose regimen for anyone.

**Methods:** Plasma or serum concentrations of Vitamin D_3_ and 25(OH)D_3_ from different trials were compiled together. We then developed a series of Physiologically-Based Pharmacokinetic (PBPK) models for vitamin D_3_ and 25(OH)D_3_ in a stepwise manner to select the best model to optimally recapitulate the 10μg and 100μg daily dose data. Each arm of the clinical trials was simulated individually. Model predictions were qualified with PK data at other doses.

**Results:** From data exploration, we observed an interesting phenomenon: the increase in plasma 25(OH)D_3_ after repeat dosing was negatively correlated with 25(OH)D_3_ baseline levels. Our final model assumes a first-order vitamin D_3_ absorption, linear vitamin D_3_ elimination and a non-linear 25(OH)D_3_ elimination which is described with an Emax function. This model offers a simple explanation to the apparent paradox: the negative correlation might arise from the non-linear 25(OH)D_3_ elimination process. The model was also able to accurately predict plasma 25(OH)D_3_ after repeat dosing at daily doses other than 10μg and 100μg, which was reassuring.

**Conclusions:** We developed a PBPK model to recapitulate PK of plasma vitamin D_3_ and 25(OH)D_3_. A personalised vitamin D_3_ supplementation protocol requires measurement of 25(OH)D_3_ baseline levels. This should be tested in the clinics for each individual.

## 1. Introduction

Vitamin D increases intestinal calcium absorption. It plays important roles in the prevention of osteoporosis and cancer (Chandler, PD. *et al*, 2020), as well as potential roles in diabetics, autoimmune disease and COVID-19 (Benskin L. L., 2020; Jungreis, I. & Kellis, M. 2020). Unfortunately, vitamin D deficiency is common around the world. In North America, the prevalence of serum 25(OH)D < 30 nmol/L in participants increased to 10% from 1988-1994 to 2001-2006 among which the elderly, pregnant women, the black community and obese population accounted for a large proportion (Ganji, V *et al*, 2012). In Europe, extensive studies on vitamin D level indicates the prevalence of vitamin D deficiency in most countries were over 20% except some Nordic countries (Feldman, D. *et al*, 2018). Traditional diet of cod and cod liver in Nordic regions is speculated to explain such difference (Feldman, D. *et al*, 2018). UK’s National Dietary and Nutrition Survey shows higher prevalence of hypovitaminosis D (marked by serum 25(OH)D < 40nmol/L) in the north of UK than the south, and the prevalence of hypovitaminosis D in most regions are higher than 30% in spring (Hyppönen, E. *et al*, 2007). Serum 25(OH)D was poor in the elderly (Hyppönen, E. *et al*, 2007). Worryingly, low serum 25(OH)D has also been observed in adolescents in the UK (Hyppönen, E. *et al*, 2007).

Various countries have issued conflicting vitamin D guidelines. The Recommended Dietary Allowance (RDA) proposed by Institute of Medicine (IOM) in the US is 600 IU/d for 1 to 70 years old and the value increased to 800 IU/d for 71 years old and over to maintain adequate vitamin D level (≥50 nmol/L) (Ross, A. C. et *al*, 2011). Scientific Advisory Committee on Nutrition (SACN) in the UK currently recommends a Reference Nutrient Intake (RNI) for vitamin D of 400 IU per day all over the year for the general UK population, including pregnant and lactating women and people at increased risk of vitamin D deficiency, to maintain serum 25(OH)D concentration not less than 25 nmol/L (SACN, 2016).

These official guidelines provided recommendations for minimal serum 25(OH)D requirements. In contrast, the optimal vitamin D level and dose regimen necessary for disease prevention are still controversial. For daily dosing, some studies suggested at least 800IU or more were needed to maintain serum 25(OH)D ≥ 50 nmol/L (Cashman, K.D. *et al*, 2011). However, higher doses such as 1800IU or more were suggested in order to obtain optimal benefits, with 4000IU-6000IU per day being the maximum tolerated dose (Bischoff-Ferrari, H. A. *et al*, 2007; Veugelers, P. *et al*, 2015; Heaney R.P. *et al*, 2011; Kimball, S.M. *et al*, 2017). In addition, monthly administration of a large dose such as 50,000IU of vitamin D may maintain serum 25(OH)D levels above 50nmol/L (i.e. 20ng/mL) in healthy subjects (Brunel, E. *et al*, 2013).

In addition, inter-individual variability in 25(OH)D PK is high. This necessitates establishing a model that can accurately predict 25(OH)D PK for an individual. Unfortunately, this challenge was not met by mechanistic PBPK modelling that was poorly calibrated with very few data (Sawyer, M. E. *et al*, 2015). A non-linear mixed effects model was reported to have successfully recapitulated PK data of a range of doses (Ocampo-Pelland, A.S. *et al*, 2016). However, population-level predictions by that model could be improved, as predictions deviated from observations by about 50% in some cases. Moreover, from that analysis, it was unclear if factors such as BMI and season should be considered in order to accurately predict 25(OH)D PK for an individual at any given time (Ocampo-Pelland, A.S. *et al*, 2016). These motivated us to develop a novel PBPK model that aims at helping individuals to select the appropriate dose regimen they need.

Here, we first extensively surveyed the literature to compile vitamin D_3_ and 25(OH)D_3_ PK data in response to a wide range of vitamin D_3_ oral dosing regimens. This paper only considers vitamin D_3_ and does not consider vitamin D_2_. Vitamin D_3_ and 25(OH)D_3_ are abbreviated as vitamin D and 25(OH)D onwards. We observed the increase in 25(OH)D plasma concentration above baseline was negatively correlated with basal 25(OH)D plasma levels. We then constructed a series of models based on single dose PK at a range of different doses and repeat daily dosing PK at 10μg (i.e. 400IU) and 100μg (4000IU). The best model was selected considering the goodness of fit and parametric inference. This model provided a possible explanation to the paradoxical observation and generated accurate predictions for daily dosing test set corresponding to doses not included in the training set. We propose using this PBPK model together with an assay to determine pre-dose 25(OH)D baseline levels in order to personalise dosing regimen and effectively address vitamin D deficiency. This model would also assist in the selection of post-dose follow-up date to ensure effectiveness of vitamin D dosing.

## 2. Results

### 2.1 PK data compilation and exploration

A total of 84 mean vitamin D_3_ (abbreviated as vitamin D afterwards) plasma / serum concentration time points measured in 307 subjects from 13 clinical trials were included in the vitamin D PK dataset. Among the 13 trials, 5 trials assessed single-dose administration and 10 trials assessed repeat dose administration. All but 2 articles were published after the year 2000. Daily dose ranged between 70μg (i.e. 2800IU) and 2500μg (i.e. 100000IU). Several types of dosage were used, including tablets, capsules, powders and 2 unknown cases. Most subjects were between 30 to 50 years old, including both males and females.

For 25(OH)D, 451 mean plasma concentration time points measured in 6484 subjects in 126 clinical trials were included. Over 80% study arms came from randomised controlled trials (RCT). Over 70% of data were published after the year 2000. Trials covered all continents except Antarctica, with the most numbers of trial conducted in the US, the Netherlands, UK, Canada in descending order. Subjects came from all age groups. 55.6% of subjects have BMI over 25, while no subjects were reported to have BMI lower than 18. 81% of all study arms were annotated with which season the trial was performed. Various quantification methods were used, including RIA, others (i.e. CHEMI), LC-MS, CPBA, HPLC in the order of popularity.

### 2.2 Data exploration

PK profiles of vitamin D and 25(OH)D under single dose and repeat dose were graphed in Figure 1. Final levels of vitamin D and 25(OH)D increased with dose (Figure 1). Vitamin D and 25(OH)D are known to have half-lives of approximately 20h and 15 days, respectively. In line with this, at 137.5μg QD dose (i.e. 5500IU), plasma / serum vitamin D reached PK steady states within 10 days (Figure 1B), while it took more than 50 days for 25(OH)D to reach PK steady state at a similar 125μg QD dose (i.e. 5000IU), (Figure 1D).

**Figure 1.**
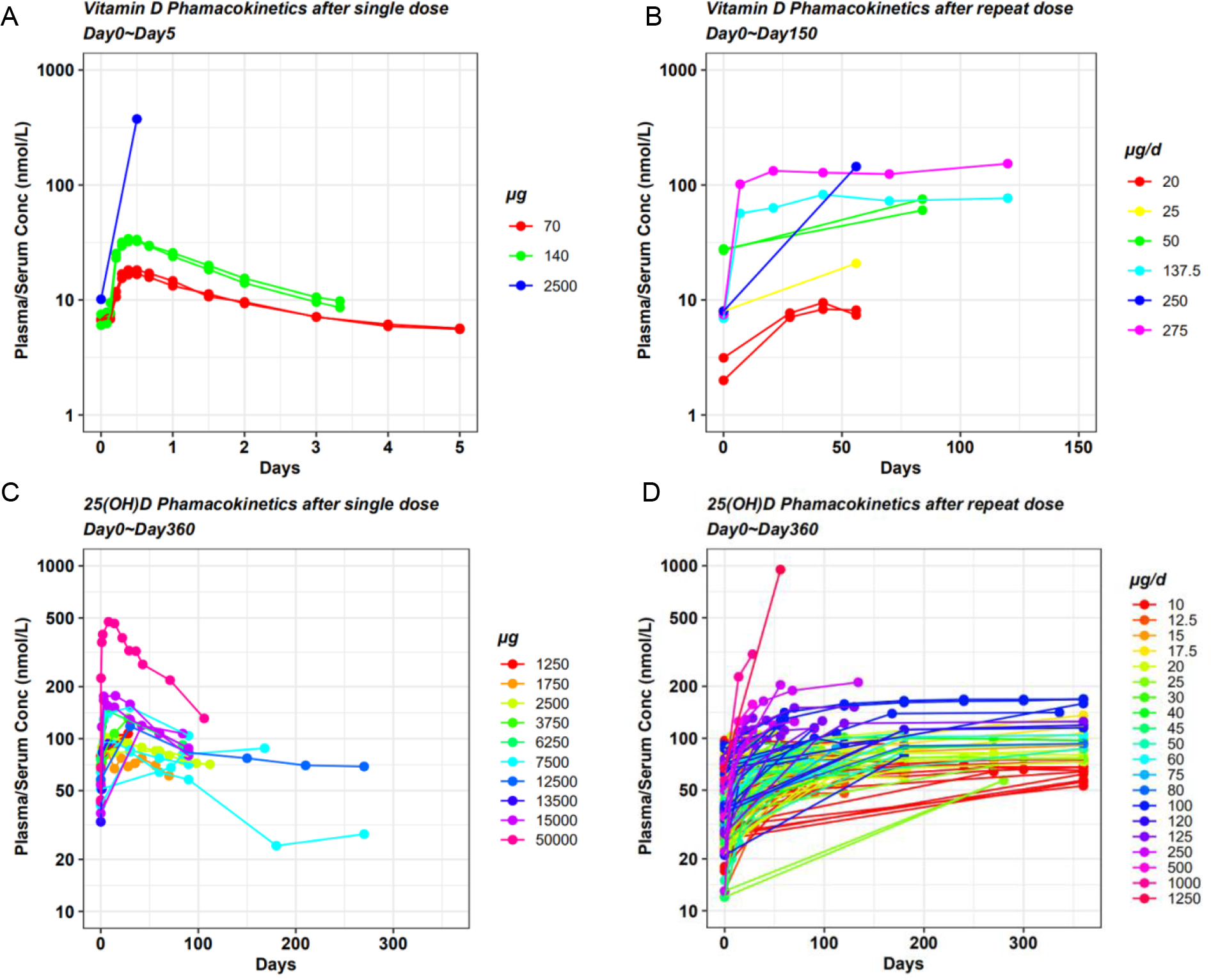
Plasma or serum PK of vitamin D and 25(OH)D after daily dosing of vitamin D across a wide range of doses. A) Vitamin D PK after single dose (day 0 - day 5). B) Vitamin D PK after repeat daily dose (day 0 – day 150). C) 25(OH)D PK after single dose (day 0 - day 360). D) 25(OH)D PK after repeat daily dose (day 0 - day 360).

We subtracted baseline 25(OH)D concentrations from their final values (taken between 14 to 720 days) to calculate increase in 25(OH)D levels (*Figure 2*A). The increase appeared constant for daily doses less than 50μg. At doses higher than 125μg, the increases were significantly larger (Figure 2A).

**Figure 2.**
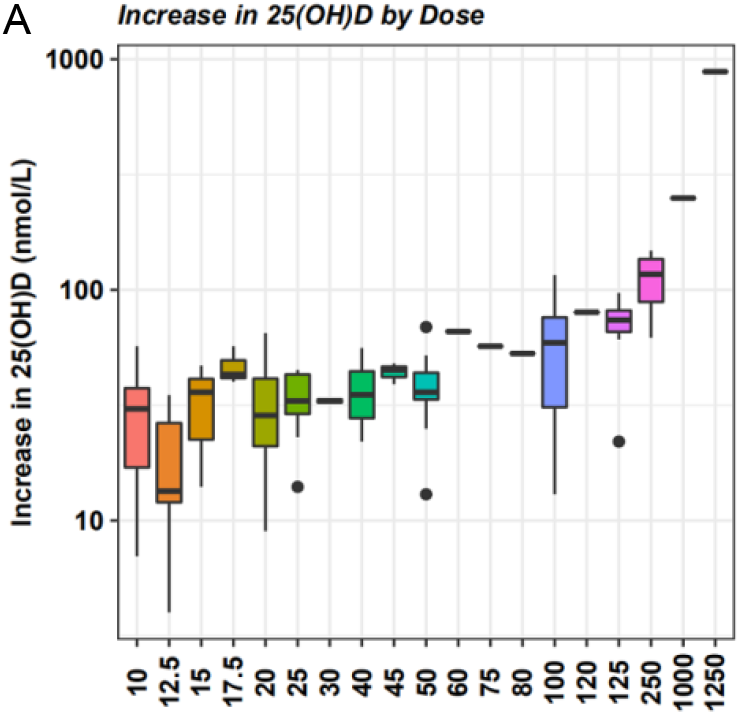
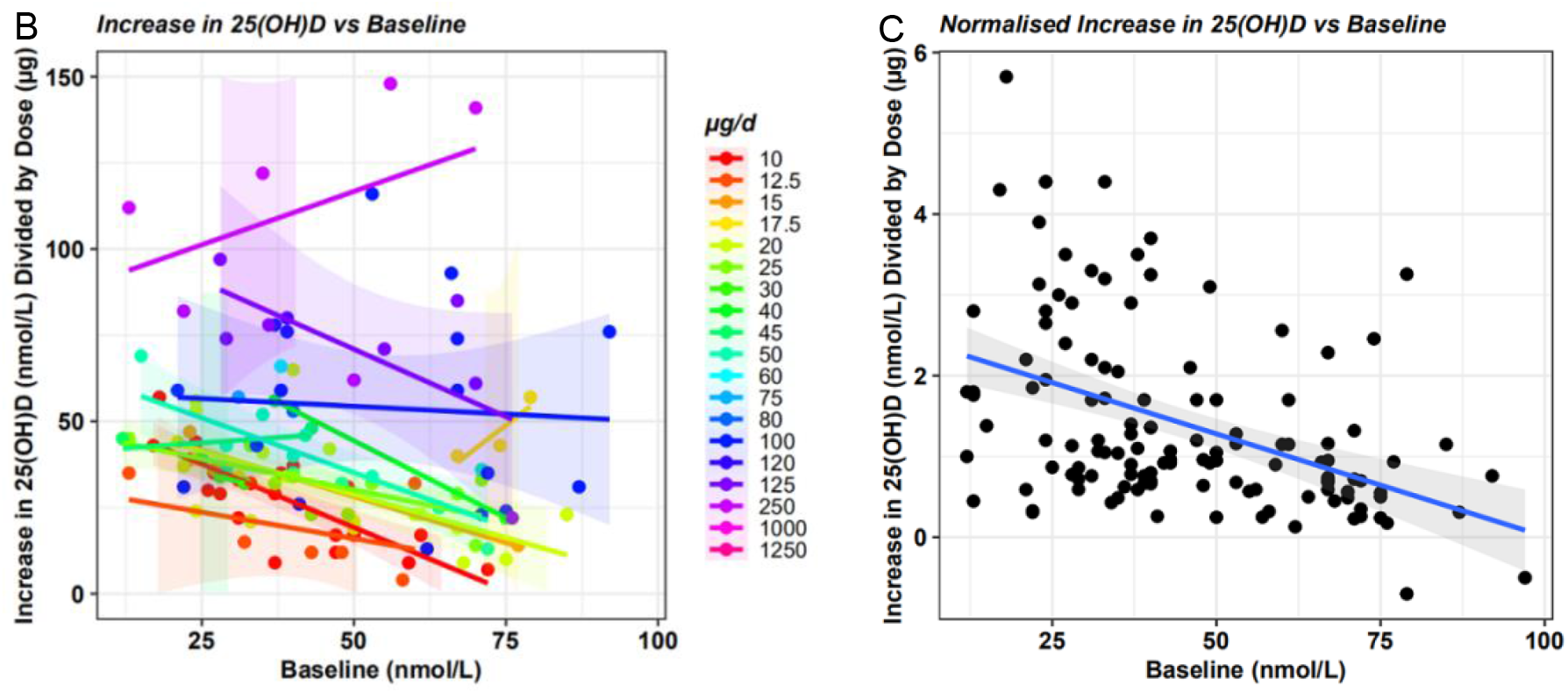
Increase in 25(OH)D plasma levels (final level – baseline level) is influenced by both dose and baseline levels. A) Change in plasma 25(OH)D concentration after repeat daily dosing is grouped by dose which ranges from 10μg (i.e. 400IU) to 1250μg (i.e. 50000IU). Duration of dosing varies between 14 to 720 days. B) Increase in plasma 25(OH)D is plotted against 25(OH)D baseline plasma levels. Each point corresponds to a study arm. C) Normalised increase in plasma 25(OH)D is plotted against 25(OH)D baseline plasma levels. Normalised increase was produced by dividing increase in plasma 25(OH)D by dose. In B and C, trend lines represent least square estimate of the means and shades mark 95% confidence intervals of the means.

We plotted the increase in 25(OH)D against baseline levels and found the two exhibited negative correlation among most dose groups (Figure 2B: daily dose ranges between 10μg to 1250μg). To remove the confounding factor dose, the increase was divided by dose (termed “normalised increase”) in Figure 2C visualise all data from Figure 2B. This analysis also revealed a negative correlation (Figure 2C).

We classified subjects into 3 groups by their plasma 25(OH)D levels, namely severe deficiency (<30 nmol/L), deficiency (30-50 nmol/L) and sufficiency (>50 nmol/L). The 30 nmol/L and 50 nmol/L cutoff values come from Food and Nutrition Board (FNB) and Institute of Medicine (IOM) (1997) and IOM’s Dietary Reference Intake (IOM, 2011), respectively. We then graphed normalised increase in different groups by different factors including trial duration, BMI, age, sex, season and the continent where the trial was conducted (Figure 3).

**Figure 3.**
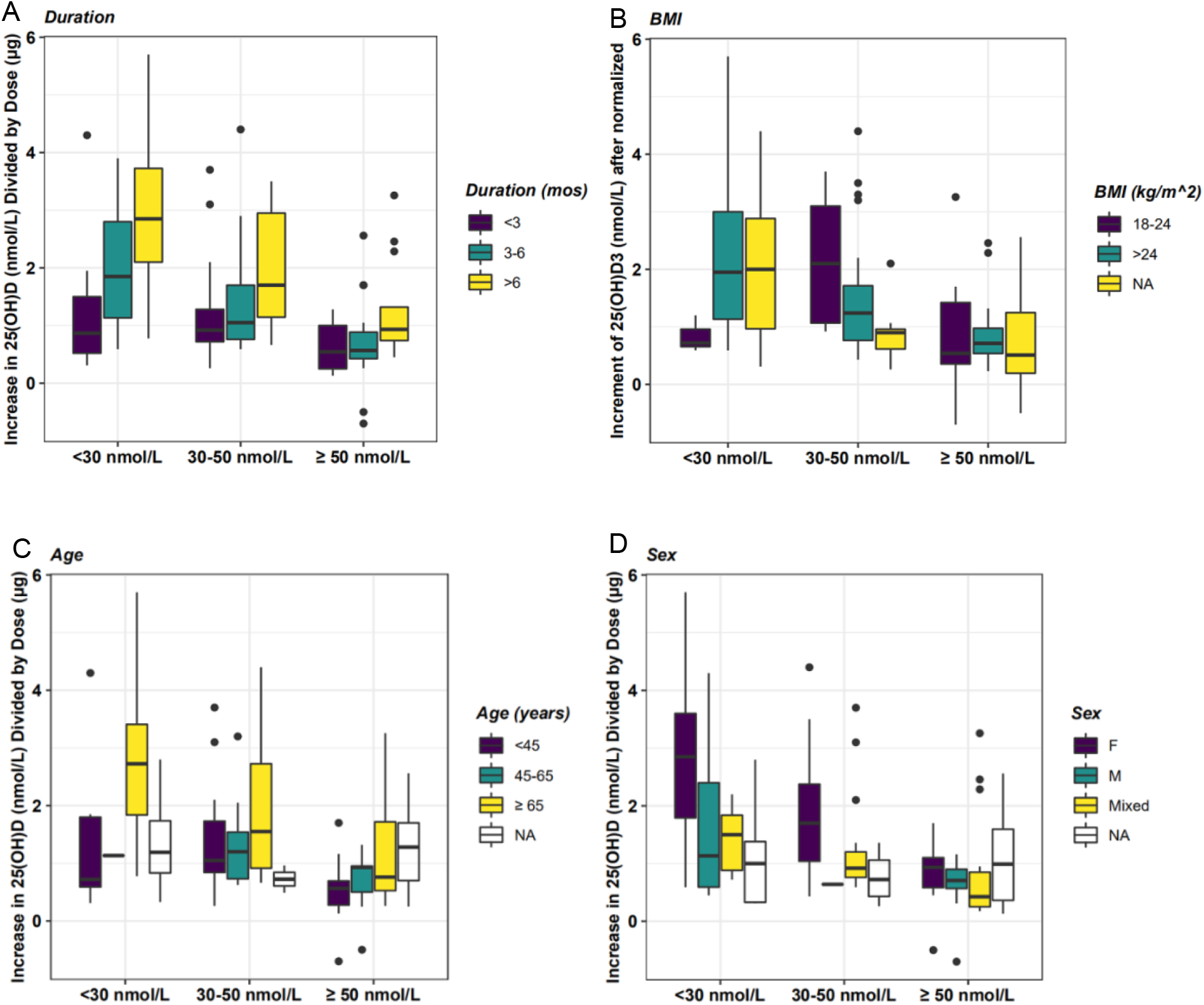
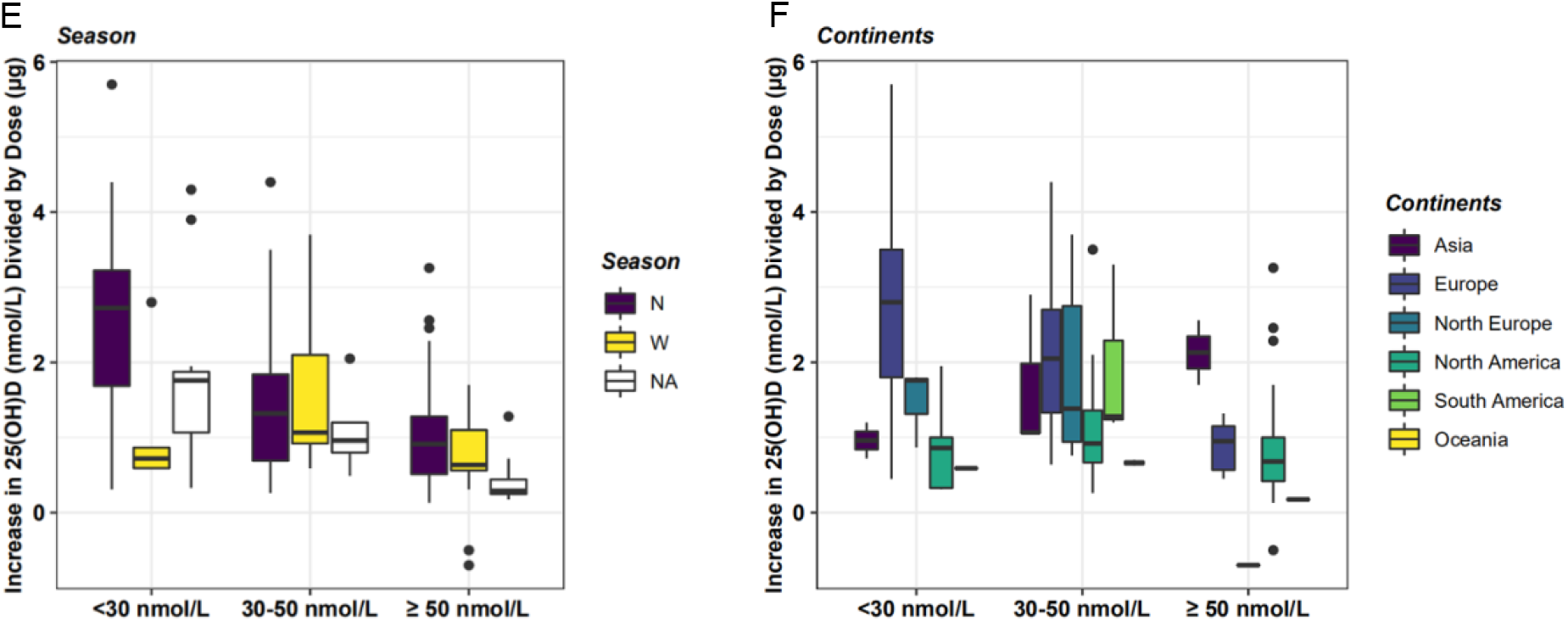
Normalised increase in 25(OH)D plasma in different baseline groups, including severe deficiency (<30 nmol/L), deficiency (30-50 nmol/L) and sufficiency (>50 nmol/L), by different factors including duration, BMI, age, sex, season and the continent where the trial took place. For season, N: non-winter; W: winter.

In the severe deficiency group and the deficiency group, normalised increase is higher at longer duration (Figure 3A). Such effect is less pronounced in the sufficiency group (Figure 3A). Similarly, female subjects exhibited higher normalised increase than male subjects in severe deficiency group and deficiency group, and the difference is smaller in the sufficiency group (Figure 3D). It is also worth noting that the normalised increase was significantly higher in winter season than non-winter season among the severely deficient (Figure 3E).

For the sake of simplicity, we next developed a model without considering any of these factors and evaluated its predictive performance in this paper.

### 2.3 PBPK Model

We developed a series of nested models (all without covariate) and selected the best model that successfully recapitulated training data and enabled good parametric inference (model evolution is described in the supplemental information). The final model considered 4 compartments, including venous, arterial, liver and the rest of body for both vitamin D and 25(OH)D (Figure 4). This model assumes a first-order absorption and a linear elimination of vitamin D, and a nonlinear 25(OH)D clearance (Figure 4). As a vitamin D dose is roughly equivalent to 1/3 dose of 25(OH)D (Shieh, A. *et al*, 2017), the rate of 25(OH)D production was assumed to be equal to 1/3 rate of vitamin D clearance. This hypothesis was verified by comparing model simulations to 7µg QD and 20µg QD repeat dose PK in the supplementary materials.

**Figure 4.**
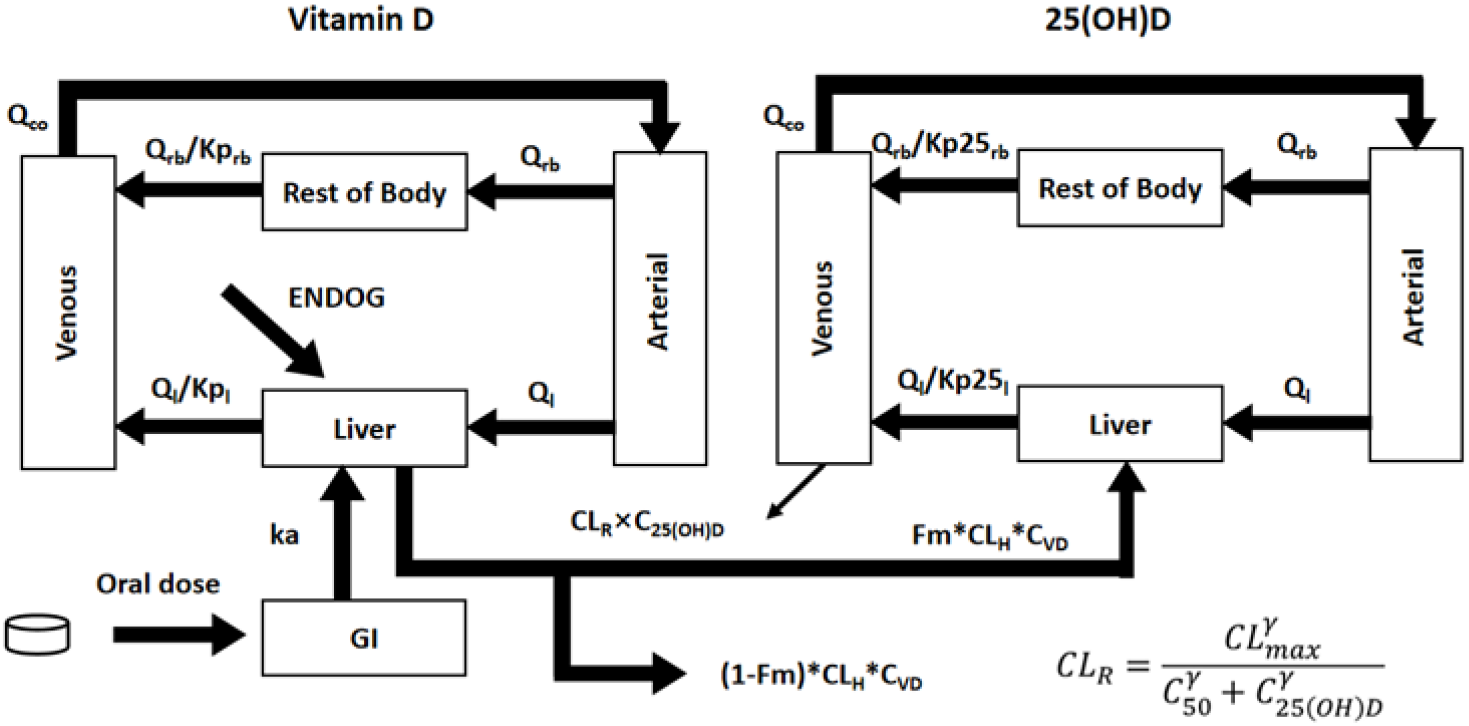
Diagram of the final model.

During parameter optimisation, the baseline level of 25(OH)D was used to estimate the initial concentrations of vitamin D and 25(OH)D in different compartments, as well as the endogenous *de novo* synthesis rate of vitamin D for any set of parameters, before simulation was carried out to evaluate objective function. In this way, observed 25(OH)D baseline level was considered for structural and parametric inference and model selection.

The prior distribution of parameters was obtained by maximum likelihood fitting to the training set (single dose: various doses; repeat dose: 10μg QD and 100μg QD), as tabulated in Table 2. We then performed MCMC simulation to obtain posterior distribution of each parameter, listed in the last column of Table 2. They exhibited low uncertainty and the expected values are similar to the prior distribution. The scatter plot between parameters showed only CL_max_ exhibited moderate degrees of correlation with *γ* and Kp25_rb_, and marginal probability distribution of each parameter has a mono-peak (Figure 5).

**Table 1.**
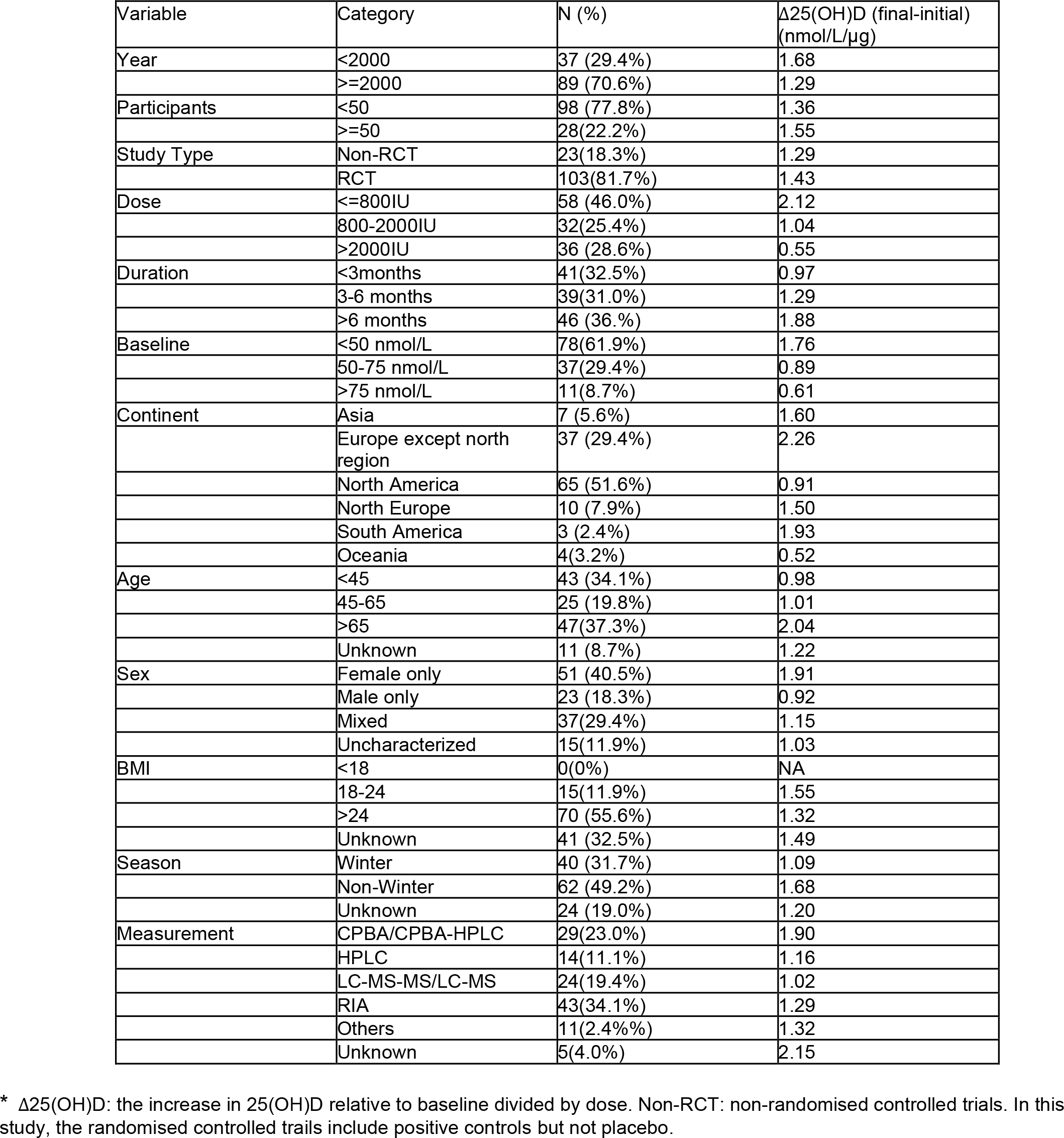
Primary characteristics of included studies in the PBPK analysis for 25(OH)D.

| Variable | Category | N (%) | Δ25(OH)D (final-initial) (nmol/L/µg) |
| --- | --- | --- | --- |
| Year | <2000 | 37 (29.4%) | 1.68 |
|  | ≥2000 | 89 (70.6%) | 1.29 |
| Participants | <50 | 98 (77.8%) | 1.36 |
|  | ≥50 | 28 (22.2%) | 1.55 |
| Study Type | Non-RCT | 23 (18.3%) | 1.29 |
|  | RCT | 103 (81.7%) | 1.43 |
| Dose | ≤800IU | 58 (46.0%) | 2.12 |
|  | 800-2000IU | 32 (25.4%) | 1.04 |
|  | >2000IU | 36 (28.6%) | 0.55 |
| Duration | <3months | 41 (32.5%) | 0.97 |
|  | 3-6 months | 39 (31.0%) | 1.29 |
|  | >6 months | 46 (36.%) | 1.88 |
| Baseline | <50 nmol/L | 78 (61.9%) | 1.76 |
|  | 50-75 nmol/L | 37 (29.4%) | 0.89 |
|  | >75 nmol/L | 11 (8.7%) | 0.61 |
| Continent | Asia | 7 (5.6%) | 1.60 |
|  | Europe except north region | 37 (29.4%) | 2.26 |
|  | North America | 65 (51.6%) | 0.91 |
|  | North Europe | 10 (7.9%) | 1.50 |
|  | South America | 3 (2.4%) | 1.93 |
|  | Oceania | 4 (3.2%) | 0.52 |
| Age | <45 | 43 (34.1%) | 0.98 |
|  | 45-65 | 25 (19.8%) | 1.01 |
|  | >65 | 47 (37.3%) | 2.04 |
|  | Unknown | 11 (8.7%) | 1.22 |
| Sex | Female only | 51 (40.5%) | 1.91 |
|  | Male only | 23 (18.3%) | 0.92 |
|  | Mixed | 37 (29.4%) | 1.15 |
|  | Uncharacterized | 15 (11.9%) | 1.03 |
| BMI | <18 | 0 (0%) | NA |
|  | 18-24 | 15 (11.9%) | 1.55 |
|  | >24 | 70 (55.6%) | 1.32 |
|  | Unknown | 41 (32.5%) | 1.49 |
| Season | Winter | 40 (31.7%) | 1.09 |
|  | Non-Winter | 62 (49.2%) | 1.68 |
|  | Unknown | 24 (19.0%) | 1.20 |
| Measurement | CPBA/CPBA-HPLC | 29 (23.0%) | 1.90 |
|  | HPLC | 14 (11.1%) | 1.16 |
|  | LC-MS-MS/LC-MS | 24 (19.4%) | 1.02 |
|  | RIA | 43 (34.1%) | 1.29 |
|  | Others | 11 (2.4%) | 1.32 |
|  | Unknown | 5 (4.0%) | 2.15 |

\* Δ25(OH)D: the increase in 25(OH)D relative to baseline divided by dose. Non-RCT: non-randomised controlled trials. In this study, the randomised controlled trials include positive controls but not placebo.

**Table 2.** Prior and posterior distributions of drug-specific parameters in the final model

| Parameter | Description | Unit | Prior distribution | Posterior distribution |
| --- | --- | --- | --- | --- |
| Vitamin D |  |  |  |  |
| Ka | Absorption rate | $\text{h}^{-1}$ | 0.09 | 0.19(0.05) |
| K<sub>pl</sub> | Partition coefficient for liver | dimensionless | 1(fixed) | NA |
| K<sub>prb</sub> | Partition coefficient for the rest of the body | dimensionless | 0.09 | 0.09(0.02) |
| SCLH | Hepatic clearance after single dose | $\text{h}^{-1}$ | 0.32 | 0.32(0.06) |
| MCLH | Hepatic clearance after multiple dose | $\text{h}^{-1}$ | 0.21 | 0.21(0.004) |
| 25(OH)D |  |  |  |  |
| F<sub>m</sub> | Fraction of Vitamin D metabolised into 25(OH)D | dimensionless | 0.33(fixed) | NA |
| Kp25<sub>rb</sub> | Partition coefficient of rest of body | dimensionless | 0.19 | 0.54 (0.15) |
| C50 | C50 in CL Emax model (CL_C50) | nmol/L | 77.9 | 86.3(4.23) |
| $\gamma$ | GAMMA in CL Emax model (CL_GAMMA) | dimensionless | 8.55 | 5.64(1.28) |
| CL<sub>max</sub> | Emax value in Emax model (CL_Emax) | L/h | 0.04 | 0.033(0.0023) |

**Figure 5.**
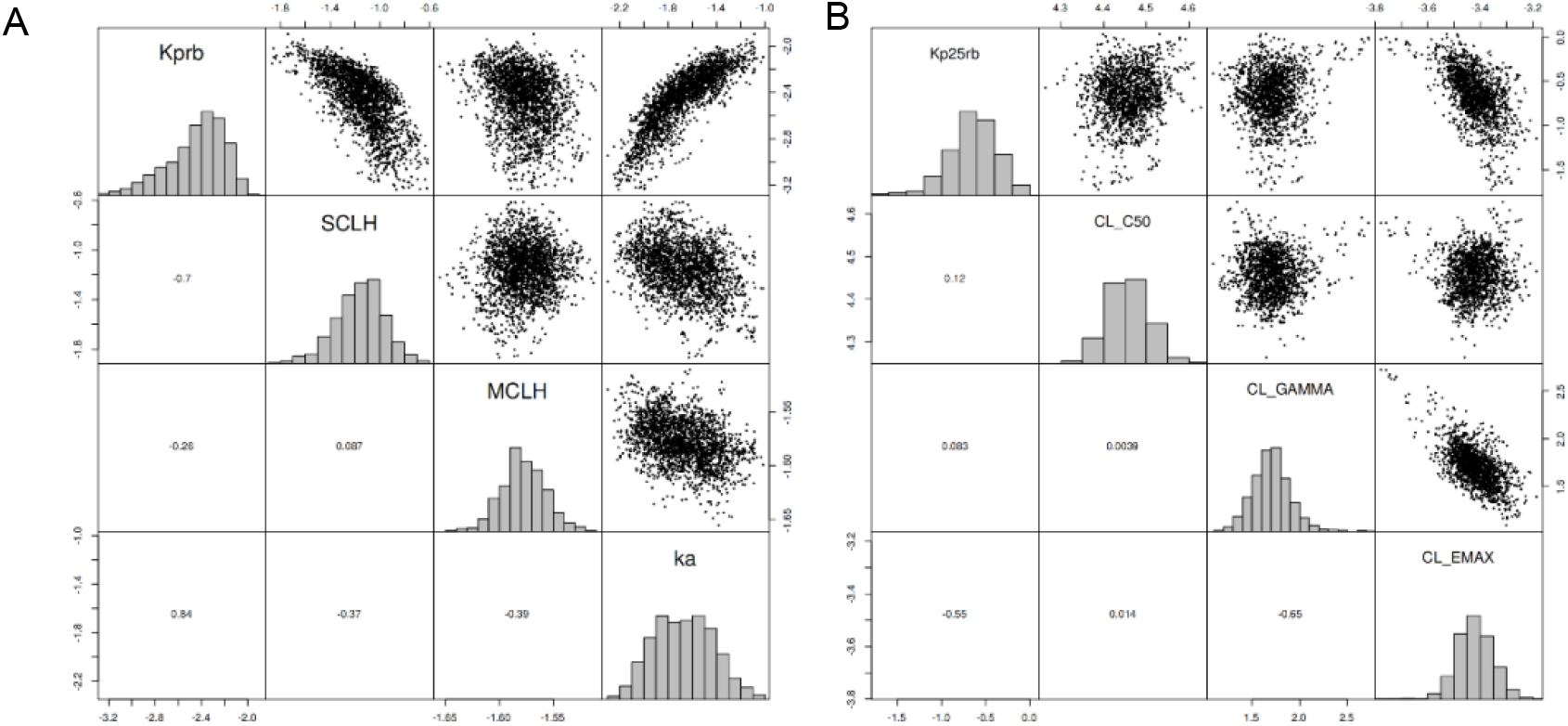
Posterior distribution of the fitted parameters from MCMC simulation for vitamin D and 25(OH)D. A) Vitamin D: 7000 iterations, 2000 burn-in. B) 25(OH)D: 3000 iterations, 1000 burn-in. Parameters were sampled and plotted in log_2_ scale.

### 2.4 Model qualification

To evaluate model fitting, we first plotted the goodness of fit for the training set. The model simulation was in good agreement with these data (Figure 6A and B). Remarkably, model predictions for the test set with daily vitamin D doses between 12.5μg and 1250μg were in good agreement with data (Figure 6C). This is excellent as the repeat doses in the test set went much beyond the range of the training set (i.e. 10μg and 100μg QD). This model also generated reasonable predictions for high vitamin D single doses between 1250μg and 50000μg, despite sometimes overestimating 25(OH)D PK for the very large doses (Figure 6D).

**Figure 6.**
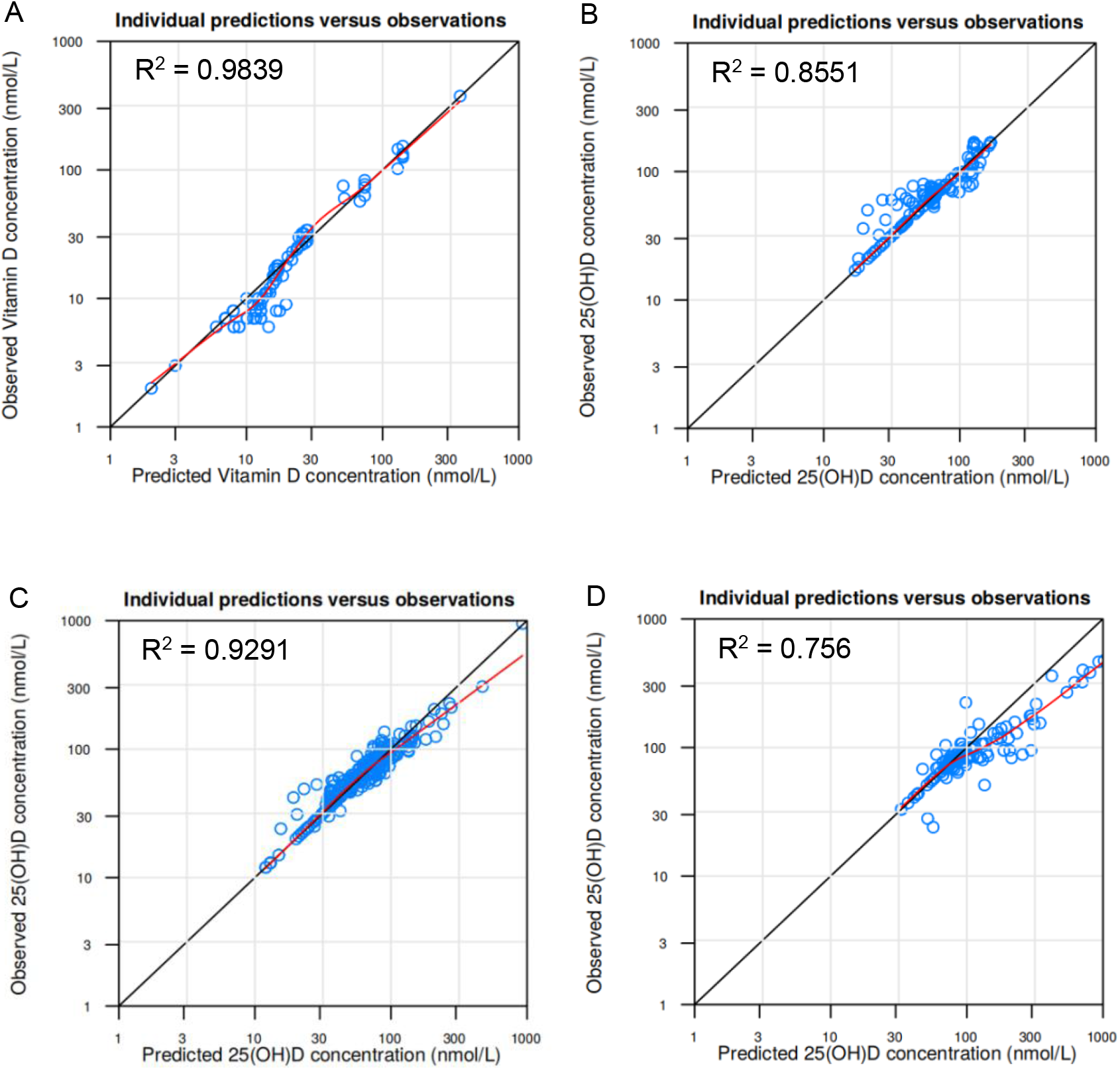
Goodness of fit plot of Vitamin D and 25(OH)D of training set and test set. A) Vitamin D after single dose (vitamin D: 70μg – 2500μg) and repeat daily dose (vitamin D: 20μg – 275μg) (training set). B) 25(OH)D at repeat vitamin D daily doses of either 10μg or 100μg (training set). C) 25(OH)D at repeat vitamin D daily doses from 12.5μg to 1250μg, except for 10μg or 100μg (test set). D. 25(OH)D at single vitamin D dose between 1250μg to 50000μg (test set). R squared was calculated in the natural scale.

The visual predictive check also illustrated model predictions were in good agreement with training set (Figure 7) and test set (supplemental material).

**Figure 7.**
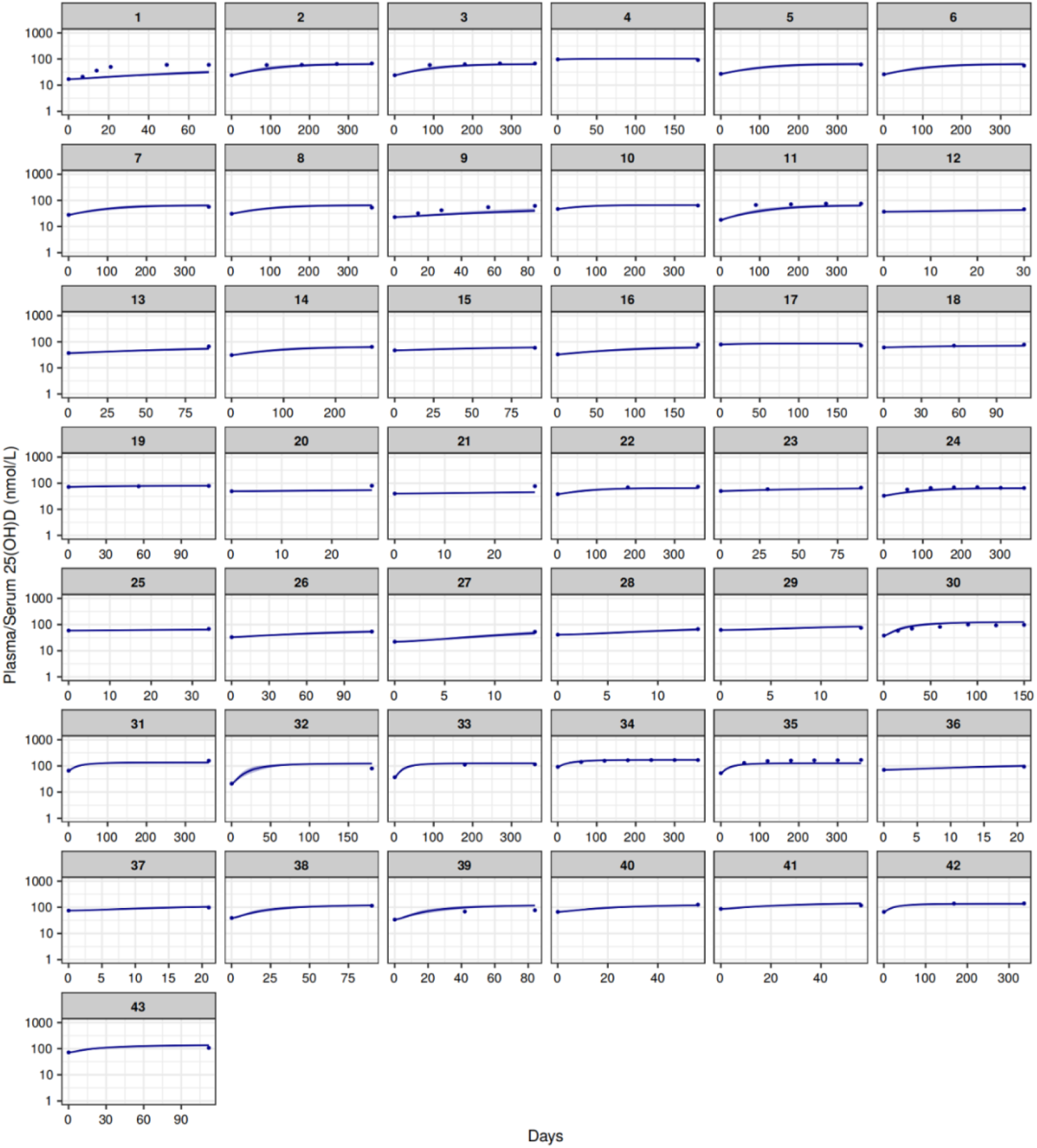
Visual predictive check for the training set. Panel 1-26: 400IU (10μg) QD; panel 27-43: 4000IU (100μg) QD.

### 2.5 Model Prediction

We then simulated 25(OH)D plasma PK under the same set of daily oral dosing regimens in two individuals with distinct 25(OH)D baseline levels to compare their responses (Figure 8). One subject has a 25(OH)D baseline at 10nmol/L (classified as severe deficiency, Figure 8A-D), and the other has a baseline at 50nmol/L (classified as insufficiency, Figure 8E-H). Interestingly, identical qualitative outcomes were predicted for both individuals at day 360: neither individual was predicted to reach the optimal threshold 75nmol/L (Holick, 2007) at 400IU QD oral dosing at day 360 (Figure 8 A and E); both were predicted to reach the optimal level or above at approximately 50% probability at 800IU QD at day 360 (Figure 8 B and F); and both were predicted to exceed 75nmol/L at 2000IU QD within 100 days (Figure 8 C and G). However, given the different baselines, the insufficient subject was predicted to reach 75nmol/L threshold quicker than the severely deficient: 130 days vs 360 days at 800IU QD; 30 days vs 75 days at 2000IU. Considering the need to ensure compliance and deliver effectiveness, 2000IU QD might be a good choice for the severely deficient subject to shorten the time to reach optimal threshold (Figure 8 C), while 800IU might be a more economical option for the sufficient subject to achieve the same goal in practice (Figure 8 F).

**Figure 8.**
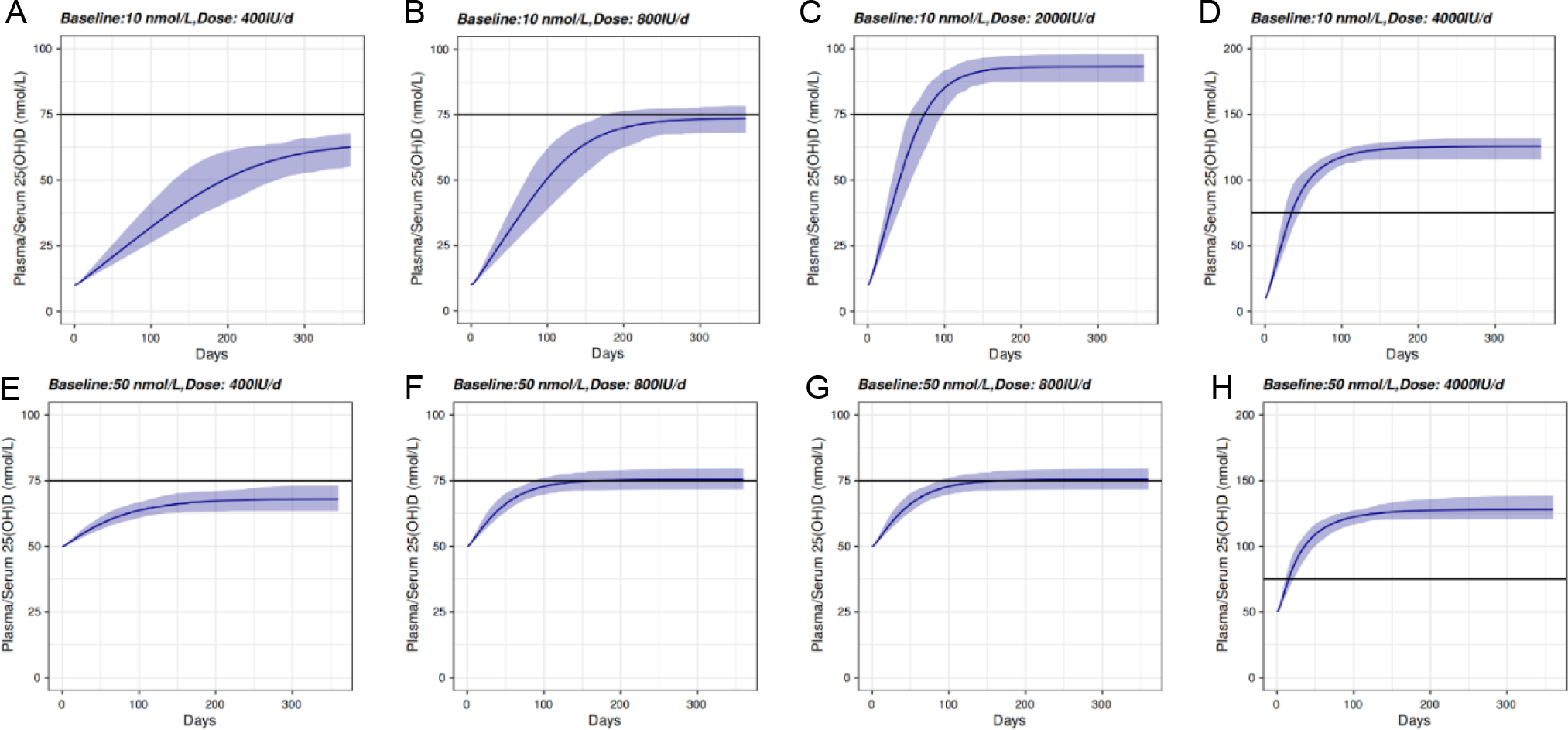
Simulation of 25(OH)D PK in two individuals with 25(OH)D baseline levels at 10 nmol/L (A-D) and 50 nmol/L (E-H). Continuous daily oral dosing at 400IU (10μg, A & E), 800IU (20μg, B & F), 2000IU (50μg, C & G) and 4000IU (100μg, D & H) were simulated for 360 days. The horizontal line marks 75 nmol/L which is regarded as the threshold for optimal plasma 25(OH)D.

We also ask the question how quickly 25(OH)D plasma levels would drop after daily oral administration is discontinued, ignoring changes in seasons and diets. Both severely deficient and sufficient subjects who undergo 800IU QD oral dosing for 180 days are predicted to exhibit similar 25(OH)D reduction profiles upon administration is discontinued after day 180 (Figure 9. Simulation of 25(OH)D PK in two individuals with 25(OH)D different baseline levels. A) Baseline = 10 nmol/L; 800IU QD; B) Baseline = 50 nmol/L, 800IU QD; C) Baseline = 10 nmol/L; 2000IU QD. D) Baseline = 50 nmol/L; 2000IU QD. The horizontal line marks 75 nmol/L regarded as the threshold for optimal plasma 25(OH)D. A and B). For the severely deficient subject who received 2000IU QD dosing for 180 days, 25(OH)D plasma concentration was predicted to decrease precipitously below 75nmol/L in about 30 days after administration was discontinued (Figure 9. Simulation of 25(OH)D PK in two individuals with 25(OH)D different baseline levels. A) Baseline = 10 nmol/L; 800IU QD; B) Baseline = 50 nmol/L, 800IU QD; C) Baseline = 10 nmol/L; 2000IU QD. D) Baseline = 50 nmol/L; 2000IU QD. The horizontal line marks 75 nmol/L regarded as the threshold for optimal plasma 25(OH)D. C). Similarly, a sufficient subject is also predicted to drop below 75nmol/L within 30 days after discontinuation (Figure 9. Simulation of 25(OH)D PK in two individuals with 25(OH)D different baseline levels. A) Baseline = 10 nmol/L; 800IU QD; B) Baseline = 50 nmol/L, 800IU QD; C) Baseline = 10 nmol/L; 2000IU QD. D) Baseline = 50 nmol/L; 2000IU QD. The horizontal line marks 75 nmol/L regarded as the threshold for optimal plasma 25(OH)D. D). Essentially, this highlights the importance of compliance in achieving and maintaining vitamin D optimum.

**Figure 9.**
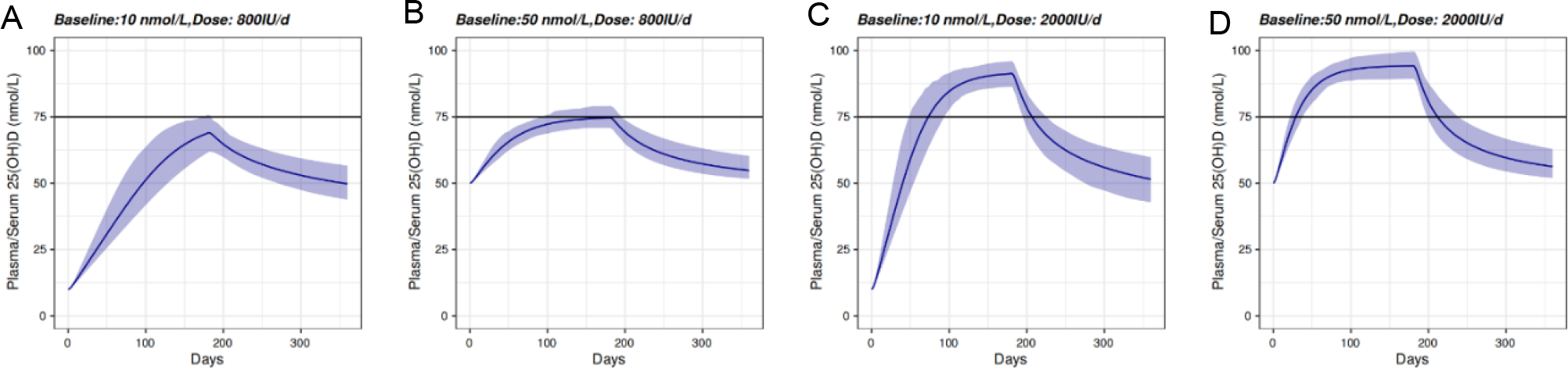
Simulation of 25(OH)D PK in two individuals with 25(OH)D different baseline levels. A) Baseline = 10 nmol/L; 800IU QD; B) Baseline = 50 nmol/L, 800IU QD; C) Baseline = 10 nmol/L; 2000IU QD. D) Baseline = 50 nmol/L; 2000IU QD. The horizontal line marks 75 nmol/L regarded as the threshold for optimal plasma 25(OH)D.

## 3. Discussions

Here, we have developed a PBPK model with a relatively simple structure, based on PK of vitamin D (single dose: 70μg, 140μg and 2500μg; repeat daily dose: 20μg – 275μg) (Figure 6A) and 25(OH)D (repeat daily dose: 10μg and 100μg) (Figure 6B). Remarkably, the model accurately predicted 25(OH)D PK in response to daily dosing between 12.5μg and 1250μg (Figure 6C) and made reasonable predictions for high single doses between 1250μg and 50000μg (Figure 6D).

Vitamin D and 25(OH)D are both lipophilic and are known to be stored in adipose tissue. However, the PK data we collected did not characterise tissue disposition (e.g. storage in adipose tissue), and BMI did not affect normalised increase in 25(OH)D in a consistent manner across 3 baseline groups (Figure 3B). Hence, we had to ignore adipose tissue disposition to avoid overfitting the data. This model may be further improved in the future to consider adipose tissue disposition.

An interesting discovery we made was that the increase in 25(OH)D plasma or serum concentration was negatively correlated with its baseline value prior to vitamin D dosing. This was observed when absolute increase in 25(OH)D was plotted against 25(OH)D baseline levels in different dose groups. When normalised increase (defined as absolute increase divided by dose) was plotted against baseline to remove the effect of dose, the negative correlation was still conspicuous (Figure 2).

The structure of the final model may offer an explanation to this. It assumes a linear vitamin D metabolism and a nonlinear elimination for 25(OH)D. Hence, a relatively high 25(OH)D baseline might correspond to a disproportionally high basal rate for 25(OH)D clearance. Given the mass balance, this consequently means the rate of endogenous *de novo* vitamin D synthesis is disproportionally high. Therefore, when the same vitamin D dose is applied to an individual with a relatively high 25(OH)D baseline level, the effect of vitamin D dosing would be expected to be relatively small in comparison to endogenous vitamin D synthesis rate. Consequently, this is expected to lead to a relatively small increase in 25(OH)D.

This structure also leads to another prediction: a high vitamin D dose may saturate the nonlinear 25(OH)D elimination and leads to more rapid increase in 25(OH)D (compare among Figure 8A-D and among Figure 8 E-H). In line with this, Figure 1C shows these very high oral doses may raise 25(OH)D above target levels in only a few days, in sharp contrast to the weeks it takes under doses used in Figure 1D.

To further verify the novel hypotheses of saturable 25(OH)D elimination rate and linear 25(OH)D production rate, quantifying kinetics of the enzymes related to 25(OH)D clearance and the expression levels of those enzymes may provide further evidence.

25(OH)D metabolism is known to take place in kidney. Unfortunately, we did not find sufficient data to support construction of a model considering this process. For the sake of simplicity, 25(OH)D metabolism was represented by the venous compartment in the final model instead. This serves our purpose of constructing a useful PK model to inform the choice of personalised dosing for healthy individuals. We recognise that modelling 25(OH)D metabolism in kidney may help us better explore 25(OH)D PK in individuals / populations with renal impairment in the future.

It is worth noting that healthy subjects in the data set we compiled refer to people without disease that are known to affect vitamin D PK. However, they may suffer from other types of disease, especially the elderly subjects reported in those studies. It is interesting to note that normalised increase in 25(OH)D was higher in those ≥ 65 years old than those < 45 years old, in both <30 nmol/L group and ≥ 50 nmol/L group (Figure 3C). We suspect compliance in the over 65 groups might be better but there is no evidence to support this.

## 4. Conclusions

Here, we compiled clinical plasma and serum PK profiles for vitamin D and 25(OH)D in response to a wide range of doses. We then developed a minimalist PK model to recapitulate the PK of vitamin D and its metabolite 25(OH)D from a training set data, and each parameter carried unambiguous physiological meaning. Importantly, predictions of this model were in good agreement with test set data (12.5μg – 50000μg) that were not used for structural and parametric inference. The model also embodies a novel hypothesis for why 25(OH)D baseline level might influence increase in 25(OH)D under a given dosing regimen. Our modelling also highlights the importance of measuring 25(OH)D for predicting temporal PK response in an individual. Using this model, we are able to assess if a daily dose or indeed any dose regimen is helpful for an individual to achieve optimal vitamin D (marked by 75 nmol/L serum concentration), estimate the time it takes to reach target 25(OH)D level in order to schedule follow up blood test, and handle real world situations that may involve missing doses and drug holiday. We believe putting such model-informed decision making into clinical practice would deliver better results in the clinics.

## 5. Method

The workflow of this study includes data compilation and exploration, PBPK model development, PBPK model validation, and model application (Figure 10). Details are provided in each section below.

**Figure 10.**
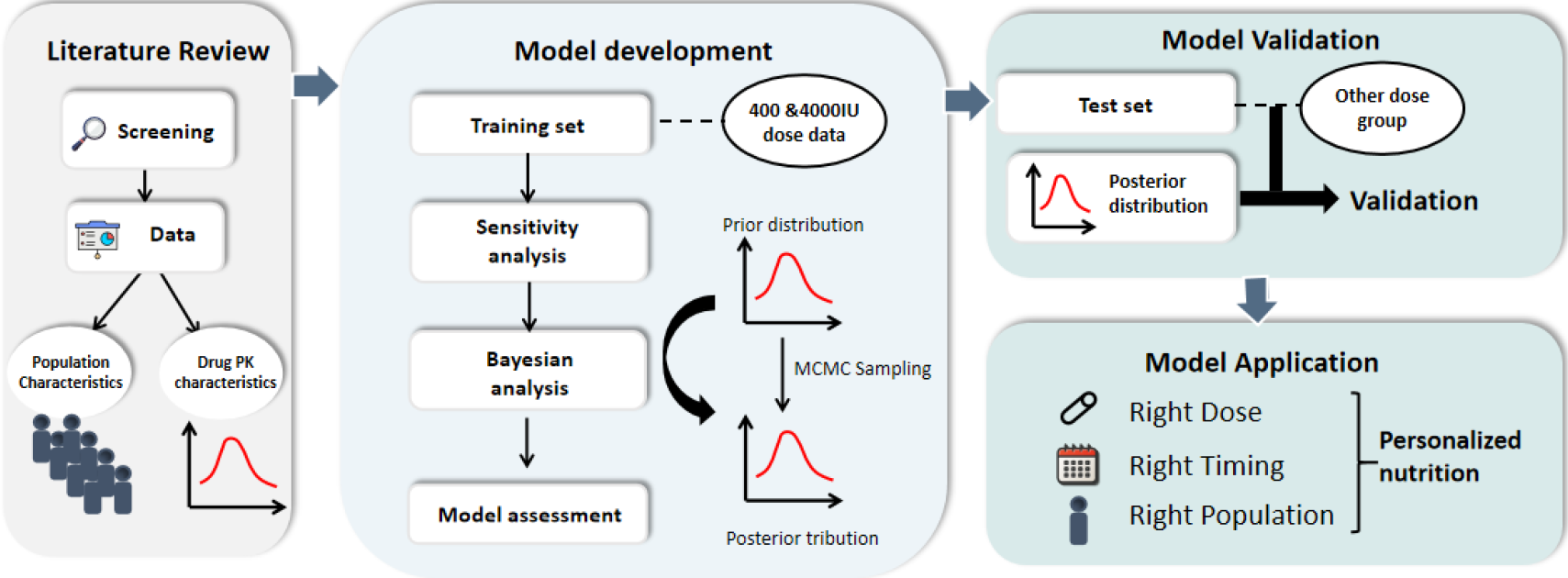
Workflow of this study includes data collection and exploration, PBPK model development, PBPK model validation, and model application.

### 5.1 Clinical data compilation and exploration

We systematically retrieved articles published in English language between January 1970 and January 2019, which are accessible by PubMed, Google Scholar and clinical trial website “https://clinicaltrials.gov/“. The keywords include vitamin D, cholecalciferol, 25-hydroxyvitamin D, 25(OH)D, plasma and pharmacokinetics. Clinical trials of vitamin D_3_ were performed using single dose and repeat daily dosing. We left out trials on dose regimens other than daily dose. Real-world evidence data were generated with either vitamin D or 25(OH)D.

We have intentionally left out all studies that were known to be conducted using vitamin D_2_, as the PK might be different from vitamin D_3_. We included those studies that did not specify if vitamin D_2_ or D_3_ was used as vitamin D_3_.

We included all subjects aged 18 and over without disease or conditions which might influence PK of vitamin D or 25(OH)D. Additionally, subjects who were prescribed with vitamin D_3_ ≥2000 IU/daily prior to study enrolment or drugs that may affect vitamin D_3_ PK during the trial were excluded. Figures were digitised using WebPlotDigitizer to obtain plasma or serum concentrations (https://automeris.io/WebPlotDigitizer/). Imputation methods for missing data were described in Supplemental Materials.

### 5.2 Model development

We first constructed a PK model for vitamin D_3_. Based on this model at nominal parameter values, we subsequently developed a 25(OH)D PK model. It was estimated that approximately 75% of vitamin D_3_ might be stored in adipose tissue. To best recapitulate plasma or serum PK data compiled for 400IU daily and 4000IU daily groups, we lumped non-elimination organs other than adipose tissue into a single compartment called “the rest of body”. The initial draft model considers 6 compartments, including adipose, liver, venous, arterial, depot and the rest of the body.

For the sake of simplicity, we assumed linear elimination of vitamin D_3_ and 25(OH)D_3_, all vitamin D_3_ was metabolised into 25(OH)D_3_, and 100% oral bioavailability. Each compartment was assumed well-stirred and the distribution was perfusion-limited.

In a non-elimination organ,

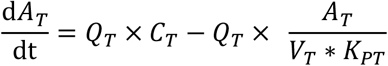

In an elimination organ,

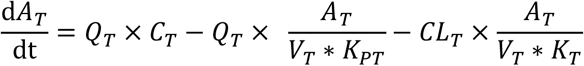

where *A*_*T*_ (nmol) is the amount of drug in tissue, *C*_*T*_ (nmol/L) is the concentration in a specific tissue, *Q*_*T*_ (L/h) is the blood flow of the tissue, *V*_*T*_ (L) is the volume of distribution for that tissue, *K*_*PT*_ (dimensionless number) is tissue to plasma partition coefficient and *CL*_*T*_ (L/h) is the plasma clearance of drug in the tissue. *K*_*PT*_ and *CL*_*T*_ may be formulated as the following:

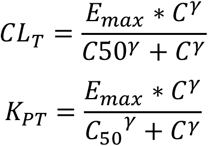

where *V*_*max*_ represents maximum reaction rate and *K*_*m*_ is the Michaelis-Menten constant, *E*_*max*_ is maximum effects, *C*_50_ is the concentration at which half of maximum effect is reached and *γ* is the exponent in the Emax model.

### 5.3 Software

The modelling work was performed in R (v3.6.0) with deSolve package (v1.28) and FME package (v1.3.5). ODE simulation was performed in Fortran 90 compiled in gfortran in RStudio Cloud environment (https://rstudio.cloud).

### 5.4 Sensitivity analysis

Local sensitivity analysis was performed with FME package. Normalised sensitivity component S_i,j_ is given by

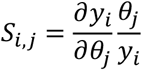

where *y*_*i*_ is an output variable and *θ*_*j*_ is a parameter.

We then computed collinearity index to evaluate identifiability for parameters in pairs or in groups. Collinearity index is defined as

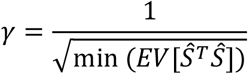

where

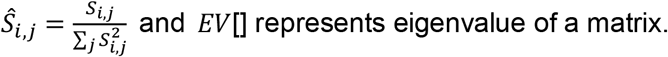

For a pair of orthogonal parameters, write down a matrix *Ŝ* of *n*x2, *n* is the number of observations. By definition, the dot product of an orthonormal matrix is the identity matrix with eigenvalue 1. This gives collinearity index *γ* = 1.

A collinearity index *γ* means that a change in the results caused by a change in one parameter can be compensated by the fraction 1 − 1/*γ* by an appropriate change of the other parameters.

A cut-off for collinearity index *γ* is often set to 10, where 90% of the parameter’s influence on the observation can be compensated by all other parameters. This way, we identified unidentifiable pairs and groups parameters which informed the choice of which parameters to keep constant and which parameters to fit.

### 5.5 Bayesian analysis

Model fitting assumes errors in plasma / serum concentration follow normal distribution, and this produces prior distributions. Functions including optim, nls, and function nlminb, from R’s base packages (R Development Core Team 2009) and the Levenberg-Marquardt algorithm from package minpack.lm (Elzhov and Mullen 2009) were used to obtain maximum likelihood estimate of parameters. We then applied the delayed rejection and adaptive Metropolis (DRAM) procedure (Haario et al. 2006; Laine 2008) to perform MCMC simulation (4000 iterations including 1000 iterations for burn-in, lower = log(exp(Prior)/10), upper = log(exp(Prior)*10), updatecov = 100, see R script for further details) and obtained posterior distribution, using FME package (modMCMC function). Visual predictive check was used to evaluate the model predictive performance in individuals.

### 5.6 Model assessment and selection

A set of nested models was developed in serial. Model with significant reduction in AIC and BIC was deemed favourable. Several diagnostic plots were evaluated:

- The dependent variable (DV) versus the individual predictions (IPRED)
- The absolute individual weighted residuals (|IWRES|) versus IPRED or time
- The conditional weighted residuals (CWRES) versus PRED or time
- A representative sample of IPRED, PRED and observations versus time (one plot per subject)
- A histogram or Quantile-Quantile (Q-Q) plot of random effects

### 5.7 Model qualification

25(OH)D_3_ plasma PK corresponding to doses other than the training set (i.e. 400IU daily and 4000IU daily) were compared with model predictions for model validation. Visual predictive check (VPC) plot and goodness of fit (GOF) plot were used to assess predictive performance. For VPC, we used the function sensRange in FME package to obtain the expected PK together with 5% and 95% quantiles.

## Data Availability

Data set we compiled are available for academic and commercial purposes. Please contact correspondence author for access.

## Contributions

Conceived the research: TY. Collected the data: ZH. Visualised and analysed the data: ZH & TY. Model development and qualification: ZH & TY. Prepared the manuscript: ZH & TY.

