## Supplemental Material for "Personalise Dose Regimen of Vitamin D_3_ Using Physiologically-Based Pharmacokinetic Modelling"

**Supplementary Tables**

**Table S1.** Primary characteristics of included vitamin D studies

| Ref No. | Study No. | Year | Dosing Regimen | No. of participants | Dosage form | Dose (IU) | Dose (ug) | Duration (d) | Age (year) | Weight (kg) | Sex (M/F) |
|---|---|---|---|---|---|---|---|---|---|---|---|
| 1 | 001 | 2011 | Single dose | 28 | Tablet | 2800 | 70 | 5 | 35.2 | 73 | 14/16 |
| 1 | 002 | 2011 | Single dose | 28 | Tablet | 2800 | 70 | 5 | 35.2 | 71 | 14/16 |
| 1 | 003 | 2011 | Single dose | 60 | Tablet | 5600 | 140 | 3.3 | 39.3 | 73 | 29/38 |
| 1 | 004 | 2011 | Single dose | 60 | Tablet | 5600 | 140 | 3.3 | 39.3 | 73 | 29/38 |
| 2 | 005 | 2016 | Single dose | 6 | Power | 100000 | 2500 | 2 | 24.9 | NA | 1/5 |
| 3 | 006 | 2002 | Multiple Dose | 13 | Tablet | 800 | 20 | 56 | 28.7 | NA | 13/0 |
| 3 | 007 | 2002 | Multiple Dose | 14 | Tablet | 800 | 20 | 56 | 72.8 | NA | 14/0 |
| 4 | 008 | 1998 | Multiple Dose | 13 | Capsule | 1000 | 25 | 56 | 28 | 81.9 | NA |
| 5 | 009 | 2019 | Multiple Dose | 24 | NA | 2000 | 50 | 84 | 40.54 | 84 | 8/16 |
| 5 | 010 | 2019 | Multiple Dose | 21 | NA | 2000 | 50 | 84 | 40.42 | 82 | 7/14 |
| 6 | 011 | 2008 | Multiple Dose | 15 | Tablet | 5500 | 137.5 | 120 | 38.7 | 84.8 | 15/0 |
| 6 | 012 | 2008 | Multiple Dose | 15 | Tablet | 11000 | 275 | 120 | 38.7 | 84.8 | 15/0 |
| 4 | 013 | 1998 | Multiple Dose | 10 | Capsule | 10000 | 250 | 56 | 28 | 81.9 | NA |

**Table S2.** Primary characteristics of included of single dose 25(OH)D studies

| Ref No. | Study No. | Year | No. of participants | Dosage form | Dose (IU) | Dose (g) | Duration (Days) | Age (year) | WT (kg) | Sex (M/F) |
|---|---|---|---|---|---|---|---|---|---|---|
| 1 | 001 | 2004 | 10 | Capsule | 50000 | 1250 | 28 | 33 | 89 | 10/0 |
| 2 | 002 | 2012 | 34 | NA | 70000 | 1750 | 70 | NA | NA | 0/34 |
| 3 | 003 | 2008 | 30 | Capsule | 100000 | 2500 | 120 | 59 | 71 | 15/21 |
| 4 | 004 | 2012 | 13 | NA | 100000 | 2500 | 90 | 75 | NA | NA |
| 5 | 005 | 2014 | 19 | NA | 150000 | 3750 | 28 | 26.11 | 64.1 | 0/19 |
| 6 | 006 | 2012 | 15 | Capsule | 250000 | 6250 | 84 | 24.9 | NA | 9/6 |
| 7 | 007 | 2008 | 8 | NA | 300000 | 7500 | 60 | 78.5 | NA | 0/8 |
| 8 | 008 | 2008 | 14 | Capsule | 300000 | 7500 | 270 | 78.9 | 62.8 | 6/8 |
| 9 | 009 | 2009 | 19 | Tablet | 300000 | 7500 | 168 | 43 | NA | 4/15 |
| 4 | 010 | 2012 | 12 | NA | 300000 | 7500 | 90 | 75 | NA | NA |
| 11 | 011 | 2008 | 19 | Tablet | 500000 | 12500 | 270 | 83 | NA | 11/8 |
| 12 | 012 | 2011 | 12 | NA | 540000 | 13500 | 7 | 61.1 | NA | 9/3 |
| 13 | 013 | 2010 | 20 | NA | 600000 | 15000 | 90 | 33.2 | NA | 0/20 |

| | | | | | | | | | | |
|---|---|---|---|---|---|---|---|---|---|---|
| 14 | 014 | 2012 | 32 | Tablet | 600000 | 15000 | 84 | 75.3 | NA | 21/11 |
| 15 | 015 | 2012 | 12 | NA | 600000 | 15000 | 90 | 76 | NA | 4/8 |
| 16 | 016 | 2012 | 2 | NA | 2000000 | 50000 | 106 | 92.5 | NA | 0/2 |

**Table S3.** Primary characteristics of included of daily repeat dose 25(OH)D studies.

| Ref No. | Study No. | Train No. | Test No. | Year | Study Type | No. of participants | Dosage form | Dose (ug) | Dose (IU) | Duration (d) | Country | Age (year) | Women (%) | Weight (kg) | BMI (kg/m$^2$) | Season after imputation | Measurement |
|---|---|---|---|---|---|---|---|---|---|---|---|---|---|---|---|---|---|
| 1 | 1 | 1 | NA | 1982 | Non-RCT | 9 | arachis oil | 10 | 400 | 70 | UK | 21.2 | 22 | 50.2 | NA | NA | HPLC |
| 2 | 2 | 2 | NA | 1988 | RCT | 25 | Tablets | 10 | 400 | 360 | Netherlands | 81 | 84 | NA | NA | Non-winter | CPBA |
| 2 | 3 | 3 | NA | 1988 | RCT | 30 | Tablets | 10 | 400 | 360 | Netherlands | 84 | 83 | NA | NA | Non-winter | CPBA |
| 3 | 4 | 4 | NA | 1991 | RCT | 124 | Tablets | 10 | 400 | 180 | USA | 61.4 | 100 | 68.5 | NA | Winter | CPBA |
| 4 | 5 | 5 | NA | 1995 | RCT | 177 | Tablets | 10 | 400 | 360 | Netherlands | 80.1 | 100 | 70.6 | 28.1 | Non-winter | CPBA-HPLC |
| 5 | 6 | 6 | NA | 1997 | RCT | 19 | Tablets | 10 | 400 | 360 | Netherlands | 77.8 | 100 | NA | 28.2 | Non-winter | CPBA-HPLC |
| 5 | 7 | 7 | NA | 1997 | RCT | 39 | Tablets | 10 | 400 | 360 | Netherlands | 78.5 | 100 | NA | 29.2 | Non-winter | CPBA-HPLC |
| 5 | 8 | 8 | NA | 1997 | RCT | 23 | Tablets | 10 | 400 | 360 | Netherlands | 77 | 100 | NA | 27.3 | Non-winter | CPBA-HPLC |
| 6 | 9 | 9 | NA | 1998 | RCT | 15 | Tablets | 10 | 400 | 90 | Netherlands | 84.4 | 100 | NA | NA | NA | RIA |
| 7 | 10 | 10 | NA | 2002 | RCT | 34 | cod liver oil | 10 | 400 | 360 | Norway | 84.4 | 74.9 | 59.4 | 22.3 | Non-winter | HPLC |
| 8 | 11 | 11 | NA | 2003 | RCT | 95 | chewable tablets | 10 | 400 | 360 | France | 74 | 100 | NA | 26.8 | Non-winter | Others |
| 9 | 12 | 12 | NA | 2003 | RCT | 67 | chewable tablets | 10 | 400 | 30 | Denmark | 75 | 60 | NA | NA | Winter | RIA |
| 10 | 13 | 13 | NA | 2005 | Non-RCT | 163 | NA | 10 | 400 | 90 | Japan | 67.3 | 98 | NA | 22 | Non-winter | RIA |
| 11 | 14 | 14 | NA | 2006 | RCT | 48 | Capsules | 10 | 400 | 270 | Chile | 77 | 100 | 66 | 30 | Non-winter | NA |
| 12 | 15 | 15 | NA | 2009 | RCT | 29 | Pills | 10 | 400 | 90 | Brazil | 61.3 | 100 | 60.3 | 26.7 | NA | RIA |
| 13 | 16 | 16 | NA | 2010 | RCT | 36 | Tablets | 10 | 400 | 180 | Netherlands | 82.4 | 100 | NA | 26.2 | Non-winter | NA |
| 14 | 17 | 17 | NA | 2010 | RCT | 80 | Capsules | 10 | 400 | 180 | Finland | 23 | 0 | 75.5 | 23.3 | Winter | Others |
| 15 | 18 | 18 | NA | 2011 | Non-RCT | 512 | Tablets | 10 | 400 | 112 | Canada | NA | NA | NA | NA | Winter | RIA |
| 15 | 19 | 19 | NA | 2011 | Non-RCT | 169 | Tablets | 10 | 400 | 112 | Canada | NA | NA | NA | NA | Winter | RIA |

| | | | | | | | | | | | | | | | | | |
|---|---|---|---|---|---|---|---|---|---|---|---|---|---|---|---|---|---|
| 16 | 20 | 20 | NA | 2012 | RCT | 27 | Fish oil Capsules | 10 | 400 | 28 | Norway | 28 | 63.6 | NA | 23.7 | Winter | RIA |
| 16 | 21 | 21 | NA | 2012 | Non-RCT | 28 | Multivitamin tablets | 10 | 400 | 28 | Norway | 28 | 63.6 | NA | 23.7 | Winter | RIA |
| 17 | 22 | 22 | NA | 2012 | RCT | 20 | Capsules | 10 | 400 | 360 | USA | 68 | 100 | 78 | 30.3 | Non-winter | RIA |
| 18 | 23 | 23 | NA | 2013 | RCT | 20 | Tablets | 10 | 400 | 90 | Thailand | 36 | 85 | 56.9 | 22.4 | Non-winter | LC-MS |
| 19 | 24 | 24 | NA | 2013 | RCT | 84 | Capsules | 10 | 400 | 360 | UK | 64.2 | 100 | 68.1 | 25.3 | Winter | LC-MS |
| 20 | 25 | 25 | NA | 2013 | RCT | 20 | Liquid | 10 | 400 | 34 | Canada | 58.9 | 0 | NA | 27.9 | Non-winter | LC-MS |
| 21 | 26 | 26 | NA | 2015 | RCT | 15 | Capsules | 10 | 400 | 112 | China | 35.3 | 60 | NA | 21.4 | Winter | LC-MS |
| 22 | 27 | NA | 1 | 1977 | Non-RCT | 18 | oil droplet | 12.5 | 500 | 14 | Canada | 83.4 | 100 | NA | NA | NA | CPBA |
| 22 | 28 | NA | 2 | 1977 | Non-RCT | 6 | oil droplet | 12.5 | 500 | 14 | Canada | 35 | 50 | NA | NA | NA | CPBA |
| 23 | 29 | NA | 3 | 1977 | RCT | 8 | Tablets | 12.5 | 500 | 120 | UK | NA | NA | NA | NA | Winter | CPBA |
| 23 | 30 | NA | 4 | 1977 | RCT | 8 | Tablets | 12.5 | 500 | 120 | UK | NA | NA | NA | NA | Winter | CPBA |
| 24 | 31 | NA | 5 | 2008 | RCT | 18 | Capsules | 12.5 | 500 | 77 | USA | 35.5 | 72.2 | NA | 31.7 | Winter | LC-MS |
| 25 | 32 | NA | 6 | 2013 | RCT | 24 | Tablets | 12.5 | 500 | 90 | India | NA | NA | NA | NA | Non-winter | NA |
| 26 | 33 | NA | 7 | 2007 | RCT | 55 | Tablets | 15 | 600 | 120 | Netherlands | 84.3 | 83.6 | NA | NA | Non-winter | RIA |
| 27 | 34 | NA | 8 | 2009 | RCT | 24 | Solution | 15 | 600 | 360 | Canada | 54.5 | 100 | NA | 26 | Non-winter | RIA |
| 28 | 35 | NA | 9 | 2015 | RCT | 22 | Tablets | 15 | 600 | 120 | Netherlands | 82 | 0 | NA | NA | Non-winter | RIA |
| 29 | 36 | NA | 10 | 2008 | RCT | 47 | Tablets | 17.5 | 700 | 360 | USA | 72.4 | NA | 62.6 | 23 | Non-winter | CPBA |
| 29 | 37 | NA | 11 | 2008 | RCT | 59 | Tablets | 17.5 | 700 | 360 | USA | 70 | NA | 79 | 27.3 | Non-winter | CPBA |
| 29 | 38 | NA | 12 | 2008 | RCT | 26 | Tablets | 17.5 | 700 | 360 | USA | 70.5 | NA | 89.4 | 33.1 | Non-winter | CPBA |
| 2 | 39 | NA | 13 | 1988 | RCT | 25 | Tablets | 20 | 800 | 360 | Netherlands | 81 | 88 | NA | NA | Non-winter | CPBA |
| 2 | 40 | NA | 14 | 1988 | RCT | 30 | Tablets | 20 | 800 | 360 | Netherlands | 84 | 83 | NA | NA | Non-winter | CPBA |
| 30 | 41 | NA | 15 | 1992 | RCT | 1634 | Pills | 20 | 800 | 540 | France | 84 | 100 | 56 | 24 | Non-winter | CPBA |
| 31 | 42 | NA | 16 | 1996 | Non-RCT | 41 | Capsules | 20 | 800 | 63 | Netherlands | 75.3 | 100 | NA | NA | Winter | CPBA |

| | | | | | | | | | | | | | | | | | |
|---|---|---|---|---|---|---|---|---|---|---|---|---|---|---|---|---|---|
| 31 | 43 | NA | 17 | 1996 | Non-RCT | 6 | Capsules | 20 | 800 | 28 | Netherlands | 30.1 | 100 | NA | NA | Winter | CPBA |
| 32 | 44 | NA | 18 | 1999 | Non-RCT | 10 | Tablets | 20 | 800 | 84 | USA | 70 | 100 | 69.7 | 27.4 | NA | Others |
| 33 | 45 | NA | 19 | 2000 | RCT | 79 | Tablets | 25 | 800 | 720 | UK | 59.2 | 100 | 62.4 | 24.1 | Non-winter | RIA |
| 34 | 46 | NA | 20 | 2001 | RCT | 35 | NA | 20 | 800 | 360 | UK | 45.3 | 100 | 67.6 | 25.1 | Non-winter | RIA |
| 34 | 47 | NA | 21 | 2001 | RCT | 35 | NA | 20 | 800 | 360 | UK | 49.1 | 100 | 67.4 | 25 | Non-winter | RIA |
| 35 | 48 | NA | 22 | 2002 | RCT | 13 | Tablets | 20 | 800 | 56 | USA | 28.7 | 0 | NA | 25 | Winter | CPBA |
| 35 | 49 | NA | 23 | 2002 | RCT | 14 | Tablets | 20 | 800 | 56 | USA | 72.8 | 0 | NA | 29 | Winter | CPBA |
| 36 | 50 | NA | 24 | 2003 | RCT | 62 | Tablets | 20 | 800 | 90 | Switzerland | 84.9 | 100 | 60.5 | 24.7 | Winter | RIA |
| 37 | 51 | NA | 25 | 2007 | RCT | 104 | Capsules | 20 | 800 | 90 | USA | 59.9 | 100 | 78 | 29 | Non-winter | RIA |
| 38 | 52 | NA | 26 | 2009 | RCT | 32 | Gelatinous form | 20 | 800 | 30 | Japan | 83.8 | 75 | 45 | 20.2 | Non-winter | RIA |
| 39 | 53 | NA | 27 | 2010 | RCT | 72 | Capsules | 20 | 800 | 180 | Netherlands | 40.5 | 75 | NA | 28.9 | Non-winter | RIA |
| 40 | 54 | NA | 28 | 2011 | RCT | 10 | Capsules | 20 | 800 | 120 | Switzerland | 63.45 | 100 | NA | 25.49 | NA | LC-MS |
| 17 | 55 | NA | 29 | 2012 | RCT | 21 | Capsules | 20 | 800 | 360 | USA | 68 | 100 | 75 | 28.2 | Non-winter | RIA |
| 41 | 56 | NA | 30 | 2012 | RCT | 13 | Capsules | 20 | 800 | 70 | Ireland | 57.2 | 61.5 | 79 | 28.3 | Winter | ELISA |
| 21 | 57 | NA | 31 | 2015 | RCT | 15 | Capsules | 20 | 800 | 112 | China | 31.8 | 53 | NA | 22.8 | Winter | LC-MS |
| 42 | 58 | NA | 32 | 2015 | RCT | 14 | Tablets | 20 | 800 | 182 | Netherlands | 85.5 | 57 | 69 | Normal | Non-winter | CHEMI |
| 43 | 59 | NA | 33 | 2016 | RCT | 20 | Capsules | 20 | 800 | 112 | USA | 84.9 | 70 | 71.1 | 27.5 | Non-winter | LC-MS |
| 1 | 60 | NA | 34 | 1982 | Non-RCT | 9 | arachis oil | 25 | 1000 | 70 | UK | 21.4 | 22.3 | 45.9 | Low | NA | HPLC |
| 44 | 61 | NA | 35 | 1991 | RCT | 14 | NA | 25 | 1000 | 280 | Finland | 84 | 85 | NA | NA | NA | CPBA |
| 44 | 62 | NA | 36 | 1991 | RCT | 5 | NA | 25 | 1000 | 280 | Finland | 84 | 85 | NA | NA | NA | CPBA |
| 45 | 63 | NA | 37 | 1998 | RCT | 13 | Capsules | 25 | 1000 | 56 | USA | 28 | 0 | 81.9 | 25.7 | Winter | HPLC |
| 46 | 64 | NA | 38 | 2001 | RCT | 33 | crystalline form | 25 | 1000 | 150 | Canada | 41.6 | 69.7 | 67.8 | NA | Winter | RIA |
| 47 | 65 | NA | 39 | 2003 | RCT | 15 | Tablets | 25 | 1000 | 160 | USA | 38.7 | 0 | 84.8 | 26.2 | Winter | HPLC |

| | | | | | | | | | | | | | | | | | |
|---|---|---|---|---|---|---|---|---|---|---|---|---|---|---|---|---|---|
| 24 | 66 | NA | 40 | 2008 | RCT | 20 | Capsules | 25 | 1000 | 77 | USA | 40 | 65 | NA | 30 | Winter | LC-MS |
| 19 | 67 | NA | 41 | 2013 | RCT | 90 | Capsules | 25 | 1000 | 360 | UK | 64.9 | 100 | 69.4 | 25.2 | Non-winter | LC-MS |
| 48 | 68 | NA | 42 | 2013 | Non-RCT | 9 | Capsules | 25 | 1000 | 77 | USA | NA | 88.9 | NA | NA | NA | LC-MS |
| 49 | 69 | NA | 43 | 2013 | RCT | 81 | Capsules | 25 | 1000 | 84 | USA | 51.1 | 72.8 | NA | 30.5 | Non-winter | RIA |
| 50 | 70 | NA | 44 | 2016 | RCT | 80 | NA | 25 | 1000 | 270 | Brazil | 58.8 | 100 | NA | 29.2 | Non-winter | HPLC |
| 28 | 71 | NA | 45 | 2015 | RCT | 16 | NA | 30 | 1200 | 120 | Netherlands | 53 | 0 | NA | 29 | NA | LC-MS |
| 21 | 72 | NA | 46 | 2015 | RCT | 14 | Capsules | 30 | 1200 | 112 | China | 34.3 | 50 | NA | 22 | Winter | LC-MS |
| 51 | 73 | NA | 47 | 2011 | RCT | 32 | Capsules | 40 | 1600 | 360 | USA | 77 | 64 | NA | 26.6 | Non-winter | HPLC |
| 17 | 74 | NA | 48 | 2012 | RCT | 20 | Capsules | 40 | 1600 | 360 | USA | 66 | 100 | 77 | 30 | Non-winter | RIA |
| 52 | 75 | NA | 49 | 1977 | Non-RCT | 18 | NA | 45 | 1800 | 28 | USA | NA | NA | NA | NA | NA | CPBA |
| 53 | 76 | NA | 50 | 1990 | RCT | 25 | Tablets | 45 | 1800 | 77 | Finland | 69.4 | 100 | 70.7 | NA | Winter | HPLC |
| 53 | 77 | NA | 51 | 1990 | RCT | 30 | Tablets | 45 | 1800 | 77 | Finland | 82.2 | 100 | 62.1 | NA | Winter | HPLC |
| 23 | 78 | NA | 52 | 1977 | Non-RCT | 9 | Tablets | 50 | 2000 | 180 | UK | NA | NA | NA | NA | NA | CPBA |
| 54 | 79 | NA | 53 | 1990 | RCT | 30 | Capsules | 50 | 2000 | 42 | USA | 81.1 | 83 | NA | NA | NA | CPBA |
| 55 | 80 | NA | 54 | 2009 | RCT | 78 | Tablets | 50 | 2000 | 84 | USA | 59.3 | 78.2 | NA | 26.1 | Winter | RIA |
| 56 | 81 | NA | 55 | 2011 | RCT | 17 | Capsules | 50 | 2000 | 180 | USA | 79.7 | 0 | NA | 26.7 | NA | RIA |
| 57 | 82 | NA | 56 | 2012 | RCT | 11 | Tablets | 50 | 2000 | 90 | Australia | 45.5 | 73 | 69.7 | 27 | Non-winter | CHEMI |
| 58 | 83 | NA | 57 | 2013 | RCT | 42 | Tablets | 50 | 2000 | 56 | Germany | 35.6 | 62 | NA | 24 | Winter | LC-MS |
| 49 | 84 | NA | 58 | 2013 | RCT | 83 | Capsules | 50 | 2000 | 90 | USA | 50.3 | 66.2 | NA | 31.9 | Non-winter | RIA |
| 59 | 85 | NA | 59 | 2013 | RCT | 50 | NA | 50 | 2000 | 42 | Netherlands | 64 | 4 | NA | NA | Non-winter | LC-MS |
| 60 | 86 | NA | 60 | 2014 | RCT | 109 | Capsules | 50 | 2000 | 360 | USA | 60.3 | 100 | 87.4 | 32.3 | Non-winter | CHEMI |
| 61 | 87 | NA | 61 | 2015 | RCT | 17 | NA | 50 | 2000 | 84 | USA | 36.5 | 100 | NA | 30.4 | Non-winter | RIA |
| 21 | 88 | NA | 62 | 2015 | RCT | 15 | Capsules | 50 | 2000 | 112 | China | 33.5 | 53 | NA | 22.2 | Winter | LC-MS |
| 43 | 89 | NA | 63 | 2016 | RCT | 19 | Capsules | 50 | 2000 | 112 | USA | 85.9 | 63 | 71.1 | 27.6 | NA | LC-MS |
| 17 | 90 | NA | 64 | 2012 | RCT | 21 | Capsules | 60 | 2400 | 360 | USA | 66 | 100 | 79 | 30 | Non-winter | RIA |

| | | | | | | | | | | | | | | | | | |
|---|---|---|---|---|---|---|---|---|---|---|---|---|---|---|---|---|---|
| 62 | 91 | NA | 65 | 2017 | RCT | 16 | Capsules | 25 | 2400 | 112 | USA | 36.9 | NA | NA | 25.7 | Non-winter | CHEMI |
| 63 | 92 | NA | 66 | 2014 | RCT | 22 | Tablets | 75 | 3000 | 112 | Denmark | 42.8 | 48 | NA | 26 | Winter | CHEMI |
| 17 | 93 | NA | 67 | 2012 | RCT | 20 | Capsules | 80 | 3200 | 360 | USA | 69 | 100 | 79 | 30.2 | Non-winter | RIA |
| 64 | 94 | 27 | NA | 1998 | RCT | 19 | NA | 100 | 4000 | 14 | Canada | 38 | NA | NA | NA | Non-winter | RIA |
| 64 | 95 | 28 | NA | 1998 | RCT | 18 | NA | 100 | 4000 | 14 | Canada | 38 | NA | NA | NA | Non-winter | RIA |
| 64 | 96 | 29 | NA | 1998 | RCT | 18 | NA | 100 | 4000 | 14 | Canada | 38 | NA | NA | NA | Non-winter | RIA |
| 46 | 97 | 30 | NA | 2001 | RCT | 28 | crystalline form | 100 | 4000 | 150 | Canada | 39.9 | 64.3 | 66.4 | NA | Winter | RIA |
| 65 | 98 | 31 | NA | 2009 | RCT | 54 | Solution | 100 | 4000 | 360 | Canada | 50.4 | 85 | NA | 24.9 | Non-winter | RIA |
| 66 | 99 | 32 | NA | 2009 | RCT | 42 | Capsules | 100 | 4000 | 180 | Newsland | 41.8 | 100 | NA | 27.5 | Winter | RIA |
| 17 | 100 | 33 | NA | 2012 | RCT | 20 | Capsules | 100 | 4000 | 360 | USA | 66 | 100 | 77 | 29.7 | Non-winter | RIA |
| 67 | 101 | 34 | NA | 2012 | RCT | 35 | NA | 100 | 4000 | 360 | USA | 65.3 | 0 | NA | 28.36 | Non-winter | RIA |
| 67 | 102 | 35 | NA | 2012 | RCT | 12 | NA | 100 | 4000 | 360 | USA | 63.2 | 0 | NA | 28.4 | Non-winter | RIA |
| 68 | 103 | 36 | NA | 2013 | RCT | 14 | Capsules | 100 | 4000 | 21 | USA | 31.2 | 56.7 | NA | 24.8 | Non-winter | NA |
| 68 | 104 | 37 | NA | 2013 | RCT | 15 | Capsules | 100 | 4000 | 21 | USA | 31.9 | 53.3 | NA | 24.6 | Non-winter | NA |
| 49 | 105 | 38 | NA | 2013 | RCT | 83 | Capsules | 100 | 4000 | 180 | USA | 51.3 | 65.1 | NA | 31.4 | Non-winter | RIA |
| 61 | 106 | 39 | NA | 2015 | RCT | 18 | NA | 100 | 4000 | 84 | USA | 38.3 | 88.9 | NA | 31.4 | Non-winter | RIA |
| 69 | 107 | 40 | NA | 2015 | RCT | 14 | Capsules | 100 | 4000 | 56 | USA | 32.8 | 0 | 78.8 | 23.4 | Winter | ELISA |
| 69 | 108 | 41 | NA | 2015 | RCT | 6 | Capsules | 100 | 4000 | 56 | USA | 32.2 | 0 | 76.1 | 23.1 | Winter | ELISA |
| 70 | 109 | 42 | NA | 2015 | RCT | 79 | NA | 100 | 4000 | 336 | USA | 36 | 9 | NA | 25 | Non-winter | LC-MS |
| 43 | 110 | 43 | NA | 2016 | RCT | 20 | Capsules | 100 | 4000 | 112 | USA | 89.5 | 63 | 69.6 | 27 | NA | LC-MS |
| 17 | 111 | NA | 68 | 2012 | RCT | 20 | Capsules | 120 | 4800 | 360 | USA | 65 | 100 | 84 | 32.1 | Non-winter | RIA |
| 47 | 112 | NA | 69 | 2003 | RCT | 15 | Tablets | 125 | 5000 | 130 | USA | 38.7 | 0 | 84.8 | 26.2 | Winter | HPLC |
| 71 | 113 | NA | 70 | 2009 | Non-RCT | 45 | NA | 125 | 5000 | 360 | Romania | 71 | 62 | NA | NA | Non-winter | RIA |
| 72 | 114 | NA | 71 | 2011 | RCT | 63 | Capsules | 125 | 5000 | 42 | Australia | 21.45 | 61.9 | NA | NA | NA | LC-MS |

| | | | | | | | | | | | | | | | | | |
|---|---|---|---|---|---|---|---|---|---|---|---|---|---|---|---|---|---|
| 73 | 115 | NA | 72 | 2012 | RCT | 15 | Tablets | 125 | 5000 | 90 | Australia | 47.4 | 40 | 81 | 28 | Non-winter | RIA |
| 74 | 116 | NA | 73 | 2013 | Non-RCT | 91 | Capsules | 125 | 5000 | 56 | UK | 23 | 0 | 75 | 24 | Winter | LC-MS |
| 75 | 117 | NA | 74 | 2014 | RCT | 20 | NA | 125 | 5000 | 28 | USA | 25.8 | 100 | 63.4 | 22.8 | Non-winter | LC-MS |
| 76 | 118 | NA | 75 | 2015 | RCT | 20 | Capsules | 125 | 5000 | 98 | UK | 20.1 | 0 | 80.8 | 24.3 | Winter | LC-MS |
| 52 | 119 | NA | 76 | 1977 | Non-RCT | 18 | NA | 250 | 10000 | 28 | USA | NA | NA | NA | NA | NA | CPBA |
| 1 | 120 | NA | 77 | 1982 | Non-RCT | 8 | arachis oil | 250 | 10000 | 70 | UK | 22.5 | 25 | 45.6 | NA | NA | HPLC |
| 45 | 121 | NA | 78 | 1998 | Non-RCT | 10 | Capsules | 250 | 10000 | 56 | USA | 28 | 0 | 81.6 | 25.7 | Winter | HPLC |
| 47 | 122 | NA | 79 | 2003 | RCT | 15 | Tablets | 250 | 10000 | 130 | USA | 38.7 | 0 | 84.8 | 26.2 | Winter | HPLC |
| 77 | 123 | NA | 80 | 2011 | Non-RCT | 8 | Capsules | 250 | 10000 | 28 | USA | 47 | 62.5 | 93 | 30 | NA | LC-MS |
| 52 | 124 | NA | 81 | 1977 | Non-RCT | 18 | NA | 250 | 10000 | 28 | USA | NA | NA | NA | NA | NA | CPBA |
| 52 | 125 | NA | 82 | 1977 | Non-RCT | 18 | NA | 1000 | 40000 | 28 | USA | NA | NA | NA | NA | NA | CPBA |
| 45 | 126 | NA | 83 | 1998 | Non-RCT | 14 | Capsules | 1250 | 50000 | 56 | USA | 28 | 0 | 81.6 | 25.7 | Winter | HPLC |

***** Ref.No: reference number; Train No: training set number; Test No: test set number.
*Meausrement: ”Others” refers to cases where the type of qualification method was hard to categorise despite description in the article

**Table S4.** Physiological parameters of the base model for Vitamin $D_3$ PBPK.

| Compartment | Bloodflow(fraction) | Bloodflow(L/h) | Volume(Fraction) | Volume(L/h) | Source |
|---|---|---|---|---|---|
| Lung | 100 | 312 | 0.76 | 0.53 | Brown1997 |
| Heart | 4 | 12.48 | 0.47 | 0.33 | Brown1997 |
| Kidney | 17.5 | 54.6 | 0.44 | 0.31 | Brown1997 |
| Liver,total | 22.7 | 70.824 | 2.57 | 1.80 | Brown1997 |
| Liver,hepatic artery | 4.6 | 14.352 | - | - | Brown1997 |
| GI | 15.1 | 47.112 | 1.71 | 1.20 | adjusted |
| Spleen | 3 | 9.36 | 0.26 | 0.18 | adjusted |
| Brain | 11.4 | 35.568 | 2 | 1.40 | Brown1996 |
| Adipose | 5.2 | 16.224 | 21.42 | 14.99 | Brown1997 |
| Skin | 5.8 | 18.096 | 3.71 | 2.60 | Brown1997 |
| Muscle | 19.1 | 59.592 | 40 | 28.00 | Brown1997 |
| Bone | 4.2 | 13.104 | 14.29 | 10.00 | Brown1997 |
| Rest of body (all other organs) | 10.1 | 31.512 | 4.37 | 3.06 | Brown1997 |
| Venous | 100 | 312 | 6 | 4.20 | Brown1997 |
| Arterial | 100 | 312 | 2 | 1.40 | Brown1997 |
| splanchnic organ | 18.1 | 56.472 | na | - | Brown1997 |

* The physiological parameters describe the US population with 70kg mean body weight (Brown *et al*, 1997).

Brown, R. P., Delp, M. D., Lindstedt, S. L., Rhomberg, L. R., & Beliles, R. P. (1997). *Physiological Parameter Values for Physiologically Based Pharmacokinetic Models. Toxicology and Industrial Health, 13(4), 407–484.*

**Table S5.** Physiological parameters of the PBPK model after lumping compartments.

| **Parameters** | **Definition** | **Units** | **Value** |
|---|---|---|---|
| PBPK model which considers adipose compartment | | | |
| Qco | Cardiac output | L/h | 312.000 |
| Ql | Blood flow to liver | L/h | 70.824 |
| Qadi | Blood flow to adipose | L/h | 16.224 |
| Qrb | Blood flow to rest of body | L/h | 224.952 |
| Vven | Volume of venous compartment | L | 4.20 |
| Vl | Volume of liver compartment | L | 1.80 |
| Vadi | Volume of adipose compartment | L | 14.99 |
| Vrb | Volume of rest of body compartment | L | 47.61 |
| Vart | Volume of arterial compartment | L | 1.40 |
| PBPK model which does not consider adipose compartment | | | |
| Qco | Cardiac output | L/h | 312.000 |
| Ql | Blood flow to liver | L/h | 70.824 |
| Qrb | Blood flow to rest of body | L/h | 241.176 |
| Vven | Volume of venous compartment | L | 4.2 |
| Vl | Volume of liver compartment | L | 1.8 |
| Vrb | Volume of rest of body compartment | L | 62.60 |
| Vart | Volume of arterial compartment | L | 1.40 |

* Lumping: Assume apparent density = 1kg/L. PBPK model which considers adipose compartment: Vrb = 70L – Vadi – Vl – Vven – Vart; Qrb = Qco – Qadi– QL. PBPK model which does not consider adipose compartment: Vrb = 70L – Vl – Vven – Vart; Qrb = Qco – Ql.

**Table S6.** Model development.

| Run No | Model Structure Description | OFV (-2LL) | AIC | BIC | Comments |
|---|---|---|---|---|---|
| Models considering vitamin $D_3$ PK | | | | | |
| Run001a | A first-order absorption and first-order elimination PBPK model with five compartments: venous blood, arterial blood, adipose, liver, and rest of body. Fitted assuming sd=1 | 3727.248 | 3737.248 | 3749.402 | GOF: Good MCMC iteration plot: Bad, unstable MCMC pairs: bad |
| Run001b | A first-order absorption and elimination PBPK model with five compartments: venous blood, arterial blood, adipose, liver, and rest of body. Fitted with weighted residuals sd=conc | 634.3905 | 644.3905 | 656.5446 | GOF: Bad MCMC iteration plot: Bad, unstable MCMC pairs: bad |
| Run001c | A first-order absorption and elimination PBPK model with five compartments: venous blood, arterial blood, adipose, liver, rest of body. Fitted with weighted residuals sd=conc/sqrt(n) | 584.701 | 594.701 | 606.8551 | GOF: Bad MCMC iteration plot: Bad, unstable MCMC pairs: bad |
| Run002 | A first-order absorption and elimination PBPK model with five compartments: venous blood, arterial blood, adipose, liver, rest of body. Fitting assumes inter-occasion variability | 3530.373 | 3542.373 | 3556.958 | GOF: Good MCMC iteration plot: Bad, unstable MCMC pairs: bad |
| Run003 | A first-order absorption and elimination PBPK model with four compartments: venous blood, arterial blood, adipose, liver, rest of body. Fitting assumes inter-occasion variability. Kpl, Kprb, SCLH, MCLH were fitted. | 3531.46 | 3541.46 | 3553.614 | GOF: Good MCMC iteration plot: Bad, unstable MCMC pairs: bad |
| Run004 | A first-order absorption and elimination PBPK model with four | 3531.67 | 3539.67 | 3549.393 | GOF fit: Good |

| | | | | | |
|---|---|---|---|---|---|
| (Final model) | compartments: venous blood, arterial blood, liver, rest of body. Kpl=1. Fitting assumes inter-occasion variability: Kprb, SCLH, MCLH. | | | | MCMC iteration plot: good, stable<br>MCMC pairs: good |
| Run005 | A first-order absorption and elimination PBPK model with four compartments: venous blood, arterial blood, adipose, liver, rest of body. Kpl=1. Fitting assumes inter-occasion variability: Kprb, SCLH, MCLH. | 2884.229 | 2894.229 | 2906.383 | GOF fit: Good<br>MCMC iteration plot: Bad, not stable<br>MCMC pairs: bad |
| Models considering 25(OH)$D_3$ PK | | | | | |
| Run006 | Vitamin D part:<br>A first-order absorption and elimination PBPK model with four compartments: venous blood, arterial blood, adipose, liver, rest of body. Kpl=1. Fitting assumes inter-occasion variability: Kprb, SCLH, MCLH.<br>25(OH)D part:<br>A first-order elimination model with four compartment (venous blood, arterial blood, liver, rest of body), inter-occasion variability on clearance. The metabolism of 25(OH)D was assumed happen in venous compartment | 1318.651 | 1324.651 | 1333.23 | GOF fit: Bad<br>MCMC iteration plot: stable<br>MCMC pairs: bad |
| Run007 | Viatmin D part:<br>A first-order absorption and elimination PBPK model with four compartments: venous blood, arterial blood, adipose, liver, rest of Body). Kpl=1. Fitting assumes inter-occasion variability: Kprb, SCLH, MCLH.<br>25(OH)D part:<br>A first-order elimination model with five compartments: venous blood, arterial blood, liver, kidney, rest of body. Inter-occasion | 1318.628 | 1326.626 | 1338.065 | GOF fit: Bad<br>MCMC failed |

| | | | | | |
|---|---|---|---|---|---|
| | variability on clearance was introduced in the model. 25(OH)D metabolism was assumed to happen in kidney. | | | | |
| Run008 | Vitamin D part:<br>A first-order absorption and elimination PBPK model with four compartment including venous blood, arterial blood, adipose, liver, rest of Body) and inter-occasion variability assumed. Except Kpl=1(Fixed), Kprb, SCLH,MCLH were estimated.<br>25(OH)D part:<br>A first-order elimination model with five compartment (venous blood, arterial blood, liver, kidney, rest of body). Inter-occasion variability and Emax model on clearance were introduced in the model. The metabolism of 25(OH)D was assumed happen in kidney compartment. | 1312.87 | 1322.87 | 1337.169 | GOF fit: Bad<br>MCMC failed |
| Run009 | Vitamin D part:<br>A first-order absorption and elimination PBPK model with four compartment including venous blood, arterial blood, adipose, liver, rest of Body) and inter-occasion variability assumed. Except Kpl=1(Fixed), Kprb, SCLH,MCLH were estimated.<br>25(OH)D part:<br>A first-order elimination model with five compartment (venous blood, arterial blood, liver, rest of body). Inter-occasion variability and Emax model on clearance were introduced in the model. The metabolism of 25(OH)D was assumed happen in venous compartment | 1312.855 | 1320.855 | 1332.294 | GOF fit: Good<br>MCMC iteration plot: stable<br>MCMC pairs: good. |
| Run009b (Final model) | Vitamin D part:<br>A first-order absorption and elimination PBPK model with four | 1312.857 | 11320.857 | 1332.296 | GOF fit: Good<br>MCMC iteration plot: |

| | | | | | |
|---|---|---|---|---|---|
| | compartment including venous blood, arterial blood, adipose, liver, rest of Body) and inter-occasion variability assumed. Except Kpl=1(Fixed), Kprb, SCLH,MCLH were estimated.<br>25(OH)D part:<br>A first-order elimination model with five compartment (venous blood, arterial blood, liver, rest of body). Inter-occasion variability and Emax model on clearance were introduced in the model. The metabolism of 25(OH)D was assumed happen in venous compartment. The production rate of 25(OH)D was assumed to be 1/3 of the vitamin D metabolism rate. | | | | stable<br>MCMC pairs: good |
| Run010 | Vitamin D part:<br>A first-order absorption and elimination PBPK model with four compartments: venous blood, arterial blood, adipose, liver, rest of Body) Kpl=1. Fitting assumes inter-occasion variability: Kprb, SCLH, MCLH.<br>25(OH)D part:<br>A first-order elimination model with five compartments: venous blood, arterial blood, liver, rest of body. Inter-occasion variability and Emax model were introduced to clearance. Additionally, Emax model on Kp25rb was introduced in the model. The metabolism of 25(OH)D was assumed happen in venous compartment | 1311.68 | 1323.68 | 1340.839 | GOF fit: Not good (good in low concentrations, bad in high concentrations)<br>MCMC iteration plot: Bad not stable<br>MCMC pairs: bad |

**Table S7.** Imputation methods used to fill in missing values in Table S1.

| Covariate | Methods of imputation |
|---|---|
| Subjects | The number of subjects was assumed to be constant throughout the study. For RCTs, each arm is assumed to have equal number of subjects, if the number of subjects was not available |
| Age | No imputation was conducted for missing values |
| Sex | If a gender exceeds 80%, the group was assigned that gender. If no gender exceeds 80%, sex was mixed. No imputation was conducted for missing values. |
| Weight | No imputation was conducted for missing values |
| BMI | No imputation was conducted for missing values |
| Season | Winter: only those studies with duration shorter than 6 months and all participants entered the study in winter. Missing value: study duration is less than 6 months and entry time was not specified. Non-winter: all other conditions. |
| Measurement | No imputation was conducted for missing values |
| Dosage form | No imputation was conducted for missing values |

**Table S8.** Reference for vitamin D PK on single dose and repeat dose regimens

| Ref No. | Reference Source |
|---|---|
| 1 | Denker, A. E., Lazarus, N., Porras, A., Ramakrishnan, R., Constanzer, M., Scott, B. R., … Wagner, J. A. (2011). Bioavailability of Alendronate and Vitamin D3in an Alendronate/Vitamin D3Combination Tablet. The Journal of Clinical Pharmacology, 51(10), 1439–1448. |
| 2 | Hermes, W. A., Alvarez, J. A., Lee, M. J., Chesdachai, S., Lodin, D., Horst, R., & Tangpricha, V. (2016). Prospective, Randomized, Double-Blind, Parallel-Group, Comparative Effectiveness Clinical Trial Comparing a Powder Vehicle Compound of Vitamin D With an Oil Vehicle Compound in Adults With Cystic Fibrosis. Journal of Parenteral and Enteral Nutrition, 41(6), 952–958. |
| 3 | Harris, S. S., & Dawson-Hughes, B. (2002). Plasma Vitamin D and 25OHD Responses of Young and Old Men to Supplementation with Vitamin D3. Journal of the American College of Nutrition, 21(4), 357–362. |
| 4 | Barger-Lux, M. J., Heaney, R. P., Dowell, S., Chen, T. C., & Holick, M. F. (1998). Vitamin D and its Major Metabolites: Serum Levels after Graded Oral Dosing in Healthy Men. Osteoporosis International, 8(3), 222–230. |
| 5 | Farag, H. A. M., Hosseinzadeh-Attar, M. J., Muhammad, B. A., Esmaillzadeh, A., & Hamid el Bilbeisi, A. (2019). Effects of vitamin D supplementation along with endurance physical activity on lipid profile in metabolic syndrome patients: A randomized controlled trial. Diabetes & Metabolic Syndrome: Clinical Research & Reviews. |
| 6 | Heaney, R. P., Armas, L. A., Shary, J. R., Bell, N. H., Binkley, N., & Hollis, B. W. (2008). 25-Hydroxylation of vitamin D3: relation to circulating vitamin D3 under various input conditions. The American Journal of Clinical Nutrition, 87(6), 1738–1742. |

**Table S9.** Reference for single dose 25(OH)D PK

| Ref No. | Reference Source |
|---|---|
| 1 | Armas, L. A. G., Hollis, B. W., & Heaney, R. P. (2004). Vitamin D2Is Much Less Effective than Vitamin D3in Humans. The Journal of Clinical Endocrinology & Metabolism, 89(11), 5387–5391. |
| 2 | Roth, D. E., Al Mahmud, A., Raqib, R., Black, R. E., & Baqui, A. H. (2012). Pharmacokinetics of a single oral dose of vitamin D3 (70,000 IU) in pregnant and non-pregnant women. Nutrition Journal, 11(1). |
| 3 | Ilahi, M., Armas, L. A., & Heaney, R. P. (2008). Pharmacokinetics of a single, large dose of cholecalciferol. The American Journal of Clinical Nutrition, 87(3), 688–691. |
| 4 | Rossini, M., Adami, S., Viapiana, O., Fracassi, E., Idolazzi, L., Povino, M. R., & Gatti, D. (2012). Dose-Dependent Short-Term Effects of Single High Doses of Oral Vitamin D3 on Bone Turnover Markers. Calcified Tissue International, 91(6), 365–369. |
| 4 | Rossini, M., Adami, S., Viapiana, O., Fracassi, E., Idolazzi, L., Povino, M. R., & Gatti, D. (2012). Dose-Dependent Short-Term Effects of Single High Doses of Oral Vitamin D3 on Bone Turnover Markers. Calcified Tissue International, 91(6), 365–369. |
| 5 | Meekins, M. E., Oberhelman, S. S., Lee, B. R., Gardner, B. M., Cha, S. S., Singh, R. J., … Thacher, T. D. (2014). Pharmacokinetics of daily versus monthly vitamin D3 supplementation in non-lactating women. European Journal of Clinical Nutrition, 68(5), 632–634. |
| 6 | Grossmann, R. E., Zughaier, S. M., Kumari, M., Seydafkan, S., Lyles, R. H., Liu, S., … Tangpricha, V. (2012). Pilot study of vitamin D supplementation in adults with cystic fibrosis pulmonary exacerbation. Dermato-Endocrinology, 4(2), 191–197. |
| 7 | Romagnoli, E., Mascia, M. L., Cipriani, C., Fassino, V., Mazzei, F., D'Erasmo, E., … Minisola, S. (2008). Short and Long-Term Variations in Serum Calciotropic Hormones after a Single Very Large Dose of Ergocalciferol (Vitamin D2) or Cholecalciferol (Vitamin D3) in the Elderly. The Journal of Clinical Endocrinology & Metabolism, 93(8), 3015–3020. |
| 8 | Premaor, M. O., Scalco, R., da Silva, M. J. S., Froehlich, P. E., & Furlanetto, T. W. (2008). The effect of a single dose versus a daily dose of cholecalciferol on the serum 25-hydroxycholecalciferol and parathyroid hormone levels in the elderly with secondary hyperparathyroidism living in a low-income housing unit. Journal of Bone and Mineral Metabolism, 26(6), 603–608. |
| 9 | Leventis, P., & Kiely, P. D. W. (2009). The tolerability and biochemical effects of high-dose bolus vitamin D2 and D3 supplementation in patients with vitamin D insufficiency. Scandinavian Journal of Rheumatology, 38(2), 149–153 |

| | |
|---|---|
| 10 | Bacon, C. J., Gamble, G. D., Horne, A. M., Scott, M. A., & Reid, I. R. (2008). High-dose oral vitamin D3 supplementation in the elderly. Osteoporosis International, 20(8), 1407–1415. |
| 11 | Amrein, K., Sourij, H., Wagner, G., Holl, A., Pieber, T. R., Smolle, K. H., … Dobnig, H. (2011). Short-term effects of high-dose oral vitamin D3 in critically ill vitamin D deficient patients: a randomized, double-blind, placebo-controlled pilot study. Critical Care, 15(2), |
| 12 | Cipriani, C., Romagnoli, E., Scillitani, A., Chiodini, I., Clerico, R., Carnevale, V., … Minisola, S. (2010). Effect of a Single Oral Dose of 600,000 IU of Cholecalciferol on Serum Calciotropic Hormones in Young Subjects with Vitamin D Deficiency: A Prospective Intervention Study. The Journal of Clinical Endocrinology & Metabolism, 95(10), 4771–4777. |
| 13 | Tellioglu, A., Basaran, S., Guzel, R., & Seydaoglu, G. (2012). Efficacy and safety of high dose intramuscular or oral cholecalciferol in vitamin D deficient/insufficient elderly. Maturitas, 72(4), 332–338. |
| 14 | Rossini, M., Gatti, D., Viapiana, O., Fracassi, E., Idolazzi, L., Zanoni, S., & Adami, S. (2012). Short-Term Effects on Bone Turnover Markers of a Single High Dose of Oral Vitamin D3. The Journal of Clinical Endocrinology & Metabolism, 97(4), E622–E626. |
| 15 | Van den Ouweland, J., Fleuren, H., Drabbe, M., & Vollaard, H. (2014). Pharmacokinetics and safety issues of an accidental overdose of 2,000,000 IU of vitamin D3 in two nursing home patients: a case report. BMC Pharmacology and Toxicology, 15(1). |

**Table S10.** References for repeat dose 25(OH)D PK

| Ref No. | Reference Source |
|---|---|
| 1 | Davie, M. W. J., Lawson, D. E. M., Emberson, C., Barnes, J. L. C., Roberts, G. E., & Barnes, N. D. (1982). *Vitamin D from Skin: Contribution to Vitamin D Status Compared with Oral Vitamin D in Normal and Anticonvulsant-Treated Subjects. Clinical Science, 63(5), 461–472.* |
| 2 | LIPS, P., WIERSINGA, A., VAN GINKEL, F. C., JONGEN, M. J. M., NETELENBOS, J. C., HACKENG, W. H. L., … VAN DER VIJGH, W. J. F. (1988). The Effect of Vitamin D Supplementation on Vitamin D Status and Parathyroid Function in Elderly Subjects*. The Journal of Clinical Endocrinology & Metabolism, 67(4), 644–650. |
| 3 | Dawson-Hughes, B. (1991). *Effect of Vitamin D Supplementation on Wintertime and Overall Bone Loss in Healthy Postmenopausal Women. Annals of Internal Medicine, 115(7), 505.* |
| 4 | Ooms ME, Roos JC, Bezemer PD, Van Der Vijgh WJF, Bouter LM, Lips P (1995) Prevention of bone loss by vitamin D supplementation in elderly women: a randomized double-blind trial. J Clin Endocrinol Metab 80:1052–1058 |
| 5 | Graafmans, W. C., Lips, P., Ooms, M. E., Van Leeuwen, J. P. T. M., Pols, H. A. P., & Uitterlinden, A. G. (1997). *The Effect of Vitamin D Supplementation on the Bone Mineral Density of the Femoral Neck Is Associated with Vitamin D Receptor Genotype. Journal of Bone and Mineral Research, 12(8), 1241–1245.* |
| 6 | Chel, V. G. M., Ooms, M. E., Popp-Snijders, C., Pavel, S., Schothorst, A. A., Meulemans, C. C. E., & Lips, P. (1998). *Ultraviolet Irradiation Corrects Vitamin D Deficiency and Suppresses Secondary Hyperparathyroidism in the Elderly. Journal of Bone and Mineral Research, 13(8), 1238–1242.* |
| 7 | Meyer, H. E., Smedshaug, G. B., Kvaavik, E., Falch, J. A., Tverdal, A., & Pedersen, J. I. (2002). *Can Vitamin D Supplementation Reduce the Risk of Fracture in the Elderly? A Randomized Controlled Trial. Journal of Bone and Mineral Research, 17(4), 709–715.* |
| 8 | Grados, F., Brazier, M., Kamel, S., Duver, S., Heurtebize, N., Maamer, M., … Fardellone, P. (2003). *Effects on bone mineral density of calcium and vitamin D supplementation in elderly women with vitamin D deficiency. Joint Bone Spine, 70(3), 203–208.* |
| 9 | Larsen, E. R., Mosekilde, L., & Foldspang, A. (2003). *Vitamin D and Calcium Supplementation Prevents Osteoporotic Fractures in Elderly Community Dwelling Residents: A Pragmatic Population-Based 3-Year Intervention Study. Journal of Bone and Mineral Research, 19(3), 370–378.* |
| 10 | Matsumoto, T., Miki, T., Hagino, H., Sugimoto, T., Okamoto, S., Hirota, T., … Nakamura, T. (2005). *A New Active Vitamin D, ED-71, Increases Bone Mass in Osteoporotic Patients under Vitamin D Supplementation: A Randomized, Double-Blind, Placebo-Controlled Clinical Trial. The Journal of Clinical Endocrinology & Metabolism, 90(9), 5031–5036.* |

| | |
|---|---|
| 11 | Bunout, D., Barrera, G., Leiva, L., Gattas, V., de la Maza, M. P., Avendaño, M., & Hirsch, S. (2006). Effects of vitamin D supplementation and exercise training on physical performance in Chilean vitamin D deficient elderly subjects. Experimental Gerontology, 41(8), 746–752. doi:10.1016/j.exger.2006.05.001 |
| 12 | Pignotti, G. A. P., Genaro, P. S., Pinheiro, M. M., Szejnfeld, V. L., & Martini, L. A. (2009). *Is a lower dose of vitamin D supplementation enough to increase 25(OH)D status in a sunny country? European Journal of Nutrition, 49(5), 277–283.* |
| 13 | Janssen, H. C. J. P., Samson, M. M., & Verhaar, H. J. J. (2010). *Muscle strength and mobility in vitamin D-insufficient female geriatric patients: a randomized controlled trial on vitamin D and calcium supplementation. Aging Clinical and Experimental Research, 22(1), 78–84.* |
| 14 | Laaksi, I., Ruohola, J., Mattila, V., Auvinen, A., Ylikomi, T., & Pihlajamäki, H. (2010). Vitamin D Supplementation for the Prevention of Acute Respiratory Tract Infection: A Randomized, Double-Blinded Trial among Young Finnish Men. The Journal of Infectious Diseases, 202(5), 809–814. |
| 15 | Karaplis, A. C., Chouha, F., Djandji, M., Sampalis, J. S., & Hanley, D. A. (2011). *Vitamin D Status and Response to Daily 400 IU Vitamin D3 and Weekly Alendronate 70 mg in Men and Women with Osteoporosis. Annals of Pharmacotherapy, 45(5), 561–568.* |
| 16 | Holvik, K., Madar, A. A., Meyer, H. E., Lofthus, C. M., & Stene, L. C. (2012). *Changes in the vitamin D endocrine system and bone turnover after oral vitamin D3 supplementation in healthy adults: results of a randomised trial. BMC Endocrine Disorders, 12(1).* |
| 17 | Chausmer, A. B. (2012). *Dose Response to Vitamin D Supplementation in Postmenopausal Women. Annals of Internal Medicine, 157(5), 378.* |
| 18 | Nimitphong, H., Saetung, S., Chanprasertyotin, S., Chailurkit, L., & Ongphiphadhanakul, B. (2013). *Changes in circulating 25-hydroxyvitamin D according to vitamin D binding protein genotypes after vitamin D3 or D2supplementation. Nutrition Journal, 12(1).* |
| 19 | Macdonald, H. M., Wood, A. D., Aucott, L. S., Black, A. J., Fraser, W. D., Mavroeidi, A., … Thies, F. (2013). *Hip bone loss is attenuated with 1000 IU but not 400 IU daily vitamin D3: A 1-year double-blind RCT in postmenopausal women. Journal of Bone and Mineral Research, 28(10), 2202–2213.* |
| 20 | Wagner, D., Trudel, D., Van der Kwast, T., Nonn, L., Giangreco, A. A., Li, D., … Vieth, R. (2013). *Randomized Clinical Trial of Vitamin D3Doses on Prostatic Vitamin D Metabolite Levels and Ki67 Labeling in Prostate Cancer Patients. The Journal of Clinical Endocrinology & Metabolism, 98(4), 1498–1507.* |
| 21 | Yao, P., Lu, L., Hu, Y., Liu, G., Chen, X., Sun, L., … Lin, X. (2015). *A dose–response study of vitamin D3 supplementation in healthy Chinese: a 5-arm randomized, placebo-controlled trial. European Journal of Nutrition, 55(1), 383–392.* |
| 22 | Somerville, P. J., Lien, J. W. K., & Kaye, M. (1977). The Calcium and Vitamin D Status in An Elderly Female Population and Their Response to Administered Supplemental Vitamin D3. Journal of Gerontology, 32(6), 659–663. |
| 23 | MacLennan, W. J., & Hamilton, J. C. (1977). Vitamin D supplements and 25-hydroxy vitamin D concentrations in the elderly. BMJ, 2(6091), 859–861. doi:10.1136/bmj.2.6091.860 |

| | |
|---|---|
| 24 | Holick, M. F., Biancuzzo, R. M., Chen, T. C., Klein, E. K., Young, A., Bibuld, D., … Tannenbaum, A. D. (2008). *Vitamin D2Is as Effective as Vitamin D3in Maintaining Circulating Concentrations of 25-Hydroxyvitamin D. The Journal of Clinical Endocrinology & Metabolism, 93(3), 677–681.* |
| 25 | Hiremath, V. P., Rao, C. B., Naik, V., & Prasad, K. V. (2013). Anti-inflammatory effect of vitamin D on gingivitis: A dose-response randomised control trial. Oral Health & Preventive Dentistry, 11, 61–69. https://doi.org/10.3290/j.ohpd.a29377 |
| 26 | Chel, V., Wijnhoven, H. A. H., Smit, J. H., Ooms, M., & Lips, P. (2007). Efficacy of different doses and time intervals of oral vitamin D supplementation with or without calcium in elderly nursing home residents. Osteoporosis International, 19(5), 663–671. |
| 27 | Fu, L., Yun, F., Oczak, M., Wong, B. Y. L., Vieth, R., & Cole, D. E. C. (2009). Common genetic variants of the vitamin D binding protein (DBP) predict differences in response of serum 25-hydroxyvitamin D [25(OH)D] to vitamin D supplementation. Clinical Biochemistry, 42(10-11), 1174–1177. |
| 28 | Heijboer, A. C., Oosterwerff, M., Schroten, N. F., Eekhoff, E. M. W., Chel, V. G. M., de Boer, R. A., … Lips, P. (2015). Vitamin D supplementation and testosterone concentrations in male human subjects. Clinical Endocrinology, 83(1), 105–110. |
| 29 | Blum, M., Dallal, G. E., & Dawson-Hughes, B. (2008). Body Size and Serum 25 Hydroxy Vitamin D Response to Oral Supplements in Healthy Older Adults. Journal of the American College of Nutrition, 27(2), 274–279. |
| 30 | Chapuy, M. C., Arlot, M. E., Duboeuf, F., Brun, J., Crouzet, B., Arnaud, S., … Meunier, P. J. (1992). Vitamin D3and Calcium to Prevent Hip Fractures in Elderly Women. New England Journal of Medicine, 327(23), 1637–1642. |
| 31 | Van Der Klis, F. R. M., Jonxis, J. H. P., Van Doormaal, J. J., Sikkens, P., Saleh, A. E. C., & Muskiet, F. A. J. (1996). Changes in vitamin-D metabolites and parathyroid hormone in plasma following cholecalciferol administration to pre- and postmenopausal women in the Netherlands in early spring and to postmenopausal women in Curaçao. British Journal of Nutrition, 75(04), 637. |
| 32 | Kyriakidou-Himonas, M., Aloia, J. F., & Yeh, J. K. (1999). Vitamin D Supplementation in Postmenopausal Black Women. The Journal of Clinical Endocrinology & Metabolism, 84(11), 3988–3990. |
| 33 | Hunter D, Major P, Arden N, Swaminathan R, Andrew T, Mac Gregor AJ, Keen R, Snieder H & Spector TD (2000) A ran domized controlled trial of vitamin D supplementation onpreventing postmenopausal bone loss and modifying bone metabolism using identical twin pairs. Journal of Bone and Mineral Research 15, 276–283. |
| 34 | Patel, R., Collins, D., Bullock, S., Swaminathan, R., Blake, G. M., & Fogelman, I. (2001). The Effect of Season and Vitamin D Supplementation on Bone Mineral Density in Healthy Women: A Double-Masked Crossover Study. Osteoporosis International, 12(4), 319–325. |
| 35 | Harris, S. S., & Dawson-Hughes, B. (2002). Plasma Vitamin D and 25OHD Responses of Young and Old Men to Supplementation with Vitamin D3. Journal of the American College of Nutrition, 21(4), 357–362. |

| | |
|---|---|
| 36 | Bischoff, H. A., Stähelin, H. B., Dick, W., Akos, R., Knecht, M., Salis, C., … Conzelmann, M. (2003). Effects of Vitamin D and Calcium Supplementation on Falls: A Randomized Controlled Trial. Journal of Bone and Mineral Research, 18(2), 343–351. |
| 37 | Talwar SA, Aloia JF, Pollack S, Yeh JK. Dose response to vitamin D supplementation among postmenopausal African American women. Am J Clin Nutr. 2007;86:1657–1662. |
| 38 | KUWABARA, A., TSUGAWA, N., TANAKA, K., FUJII, M., KAWAI, N., MUKAE, S., … OKANO, T. (2009). Improvement of Vitamin D Status in Japanese Institutionalized Elderly by Supplementation with 800 IU of Vitamin D3. Journal of Nutritional Science and Vitaminology, 55(6), 453–458. doi:10.3177/jnsv.55.453 |
| 39 | Wicherts, I. S., Boeke, A. J. P., van der Meer, I. M., van Schoor, N. M., Knol, D. L., & Lips, P. (2010). Sunlight exposure or vitamin D supplementation for vitamin D-deficient non-western immigrants: a randomized clinical trial. Osteoporosis International, 22(3), 873–882. |
| 40 | Bischoff-Ferrari, H. A., Dawson-Hughes, B., Stöcklin, E., Sidelnikov, E., Willett, W. C., Edel, J. O., … Egli, A. (2011). Oral supplementation with 25(OH)D3versus vitamin D3: Effects on 25(OH)D levels, lower extremity function, blood pressure, and markers of innate immunity. Journal of Bone and Mineral Research, 27(1), 160–169. |
| 41 | Cashman, K. D., Seamans, K. M., Lucey, A. J., Stöcklin, E., Weber, P., Kiely, M., & Hill, T. R. (2012). Relative effectiveness of oral 25-hydroxyvitamin D3 and vitamin D3 in raising wintertime serum 25-hydroxyvitamin D in older adults. The American Journal of Clinical Nutrition, 95(6), 1350–1356. |
| 42 | Wijnen, H., Salemink, D., Roovers, L., Taekema, D., & de Boer, H. (2015). Vitamin D Supplementation in Nursing Home Patients: Randomized Controlled Trial of Standard Daily Dose Versus Individualized Loading Dose Regimen. Drugs & Aging, 32(5), 371–378. |
| 43 | Schwartz, J. B., Kane, L., & Bikle, D. (2016). Response of Vitamin D Concentration to Vitamin D3Administration in Older Adults without Sun Exposure: A Randomized Double-Blind Trial. Journal of the American Geriatrics Society, 64(1), 65–72. |
| 44 | Sorva, A., Risteli, J., Risteli, L., Välimäki, M., & Tilvis, R. (1991). Effects of vitamin D and calcium on markers of bone metabolism in geriatric patients with low serum 25-hydroxyvitamin D levels. Calcified Tissue International, 49(S1), S88–S89. |
| 45 | Barger-Lux, M. J., Heaney, R. P., Dowell, S., Chen, T. C., & Holick, M. F. (1998). Vitamin D and its Major Metabolites: Serum Levels after Graded Oral Dosing in Healthy Men. Osteoporosis International, 8(3), 222–230. |
| 46 | Vieth R, Chan PC, MacFarlane GD. Efficacy and safety of vitamin D3 intake exceeding the lowest observed adverse effect level. Am J Clin Nutr 2001;73:288 –94. |
| 47 | Heaney, R. P., Davies, K. M., Chen, T. C., Holick, M. F., & Barger-Lux, M. J. (2003). Human serum 25-hydroxycholecalciferol response to extended oral dosing with cholecalciferol. The American Journal of Clinical Nutrition, 77(1), 204–210. |
| 48 | Biancuzzo, R. M., Clarke, N., Reitz, R. E., Travison, T. G., & Holick, M. F. (2013). Serum Concentrations of 1,25-Dihydroxyvitamin D2and 1,25-Dihydroxyvitamin D3in Response to Vitamin D2and Vitamin D3Supplementation. The Journal of Clinical Endocrinology & Metabolism, 98(3), 973–979. |

| | |
|---|---|
| 49 | Ng, K., Scott, J. B., Drake, B. F., Chan, A. T., Hollis, B. W., Chandler, P. D., … Fuchs, C. S. (2013). Dose response to vitamin D supplementation in African Americans: results of a 4-arm, randomized, placebo-controlled trial. The American Journal of Clinical Nutrition, 99(3), 587–598. |
| 50 | Cangussu, L. M., Nahas-Neto, J., Orsatti, C. L., Poloni, P. F., Schmitt, E. B., Almeida-Filho, B., & Nahas, E. A. P. (2016). Effect of isolated vitamin D supplementation on the rate of falls and postural balance in postmenopausal women fallers. Menopause, 23(3), 267–274. |
| 51 | Binkley, N., Gemar, D., Engelke, J., Gangnon, R., Ramamurthy, R., Krueger, D., & Drezner, M. K. (2011). *Evaluation of Ergocalciferol or Cholecalciferol Dosing, 1,600 IU Daily or 50,000 IU Monthly in Older Adults. The Journal of Clinical Endocrinology & Metabolism, 96(4), 981–988.* |
| 52 | Stamp, T. C. B., Haddad, J. G., & Twigg, C. A. (1977). *COMPARISON OF ORAL 25-HYDROXYCHOLECALCIFEROL, VITAMIN D, AND ULTRAVIOLET LIGHT AS DETERMINANTS OF CIRCULATING 25-HYDROXYVITAMIN D. The Lancet, 309(8026), 1341–1343.* |
| 53 | Honkanen, R., Alhava, E., Parviainen, M., Talasniemi, S., & Mönkkönen, R. (1990). The Necessity and Safety of Calcium and Vitamin D in the Elderly. Journal of the American Geriatrics Society, 38(8), 862–866. doi:10.1111/j.1532-5415. |
| 54 | Himmelstein, S., Clemens, T. L., Rubin, A., & Lindsay, R. (1990). Vitamin D supplementation in elderly nursing home residents increases 25(OH)D but not 1,25(OH)2D. The American Journal of Clinical Nutrition, 52(4), 701–706. |
| 55 | LI-NG, M., ALOIA, J. F., POLLACK, S., CUNHA, B. A., MIKHAIL, M., YEH, J., & BERBARI, N. (2009). *A randomized controlled trial of vitamin D3 supplementation for the prevention of symptomatic upper respiratory tract infections. Epidemiology and Infection, 137(10), 1396.* |
| 56 | Cherniack, E. P., Florez, H. J., Hollis, B. W., Roos, B. A., Troen, B. R., & Levis, S. (2011). *The Response of Elderly Veterans to Daily Vitamin D3 Supplementation of 2,000 IU: A Pilot Efficacy Study. Journal of the American Geriatrics Society, 59(2), 286–290.* |
| 57 | Diamond, T., Wong, Y. K., & Golombick, T. (2012). *Effect of oral cholecalciferol 2,000 versus 5,000 IU on serum vitamin D, PTH, bone and muscle strength in patients with vitamin D deficiency. Osteoporosis International, 24(3), 1101–1105.* |
| 58 | Lehmann, U., Hirche, F., Stangl, G. I., Hinz, K., Westphal, S., & Dierkes, J. (2013). *Bioavailability of Vitamin D2and D3in Healthy Volunteers, a Randomized Placebo-Controlled Trial. The Journal of Clinical Endocrinology & Metabolism, 98(11), 4339–4345.* |
| 59 | Schroten, N. F., Ruifrok, W. P. T., Kleijn, L., Dokter, M. M., Silljé, H. H., Lambers Heerspink, H. J., … de Boer, R. A. (2013). Short-term vitamin D3 supplementation lowers plasma renin activity in patients with stable chronic heart failure: An open-label, blinded end point, randomized prospective trial (VitD-CHF trial). American Heart Journal, 166(2), 357–364.e2. |
| 60 | Mason, C., Xiao, L., Imayama, I., Duggan, C., Wang, C.-Y., Korde, L., & McTiernan, A. (2014). Vitamin D3 supplementation during weight loss: a double-blind randomized controlled trial. The American Journal of Clinical Nutrition, 99(5), 1015–1025. |

| | |
|---|---|
| 62 | Shieh, A., Ma, C., Chun, R. F., Witzel, S., Rafison, B., Contreras, H. T. M., … Adams, J. S. (2017). Effects of Cholecalciferol vs Calcifediol on Total and Free 25-Hydroxyvitamin D and Parathyroid Hormone. The Journal of Clinical Endocrinology & Metabolism, 102(4), 1133–1140. |
| 63 | Nygaard, B., Frandsen, N. E., Brandi, L., Rasmussen, K., Oestergaard, O. V., Oedum, L., … Hansen, D. (2014). Effects of High Doses of Cholecalciferol in Normal Subjects: A Randomized Double-Blinded, Placebo-Controlled Trial. PLoS ONE, 9(8), e102965. |
| 64 | Trang, H. M., Cole, D. E., Rubin, L. A., Pierratos, A., Siu, S., & Vieth, R. (1998). *Evidence that vitamin D3 increases serum 25-hydroxyvitamin D more efficiently than does vitamin D2. The American Journal of Clinical Nutrition, 68(4), 854–858.* |
| 65 | Fu, L., Yun, F., Oczak, M., Wong, B. Y. L., Vieth, R., & Cole, D. E. C. (2009). Common genetic variants of the vitamin D binding protein (DBP) predict differences in response of serum 25-hydroxyvitamin D [25(OH)D] to vitamin D supplementation. Clinical Biochemistry, 42(10-11), 1174–1177. |
| 66 | Von Hurst, P. R., Stonehouse, W., & Coad, J. (2009). Vitamin D supplementation reduces insulin resistance in South Asian women living in New Zealand who are insulin resistant and vitamin D deficient – a randomised, placebo-controlled trial. British Journal of Nutrition, 103(04), 549. |
| 67 | Garrett-Mayer, E., Wagner, C. L., Hollis, B. W., Kindy, M. S., & Gattoni-Celli, S. (2012). *Vitamin D3 supplementation (4000 IU/d for 1 y) eliminates differences in circulating 25-hydroxyvitamin D between African American and white men. The American Journal of Clinical Nutrition, 96(2), 332–336.* |
| 68 | Hata, T. R., Audish, D., Kotol, P., Coda, A., Kabigting, F., Miller, J., … Gallo, R. L. (2013). *A randomized controlled double-blind investigation of the effects of vitamin D dietary supplementation in subjects with atopic dermatitis. Journal of the European Academy of Dermatology and Venereology, 28(6), 781–789.* |
| 69 | Scholten, S., Sergeev, I., Birger, C., & Song, Q. (2015). *Effects of vitamin D and quercetin, alone and in combination, on cardiorespiratory fitness and muscle function in physically active male adults. Open Access Journal of Sports Medicine, 229.* |
| 70 | Overton ET, Chan ES, Brown TT, et al. High-dose vitamin D and calcium attenuates boneloss with ART initiation: results from ACTG A5280 Abstract 133]. Top Antivir Med. 2014;22(e-1):66-67. |
| 71 | Mocanu, V., Stitt, P. A., Costan, A. R., Voroniuc, O., Zbranca, E., Luca, V., & Vieth, R. (2009). *Long-term effects of giving nursing home residents bread fortified with 125 µg (5000 IU) vitamin D3 per daily serving. The American Journal of Clinical Nutrition, 89(4), 1132–1137.* |
| 72 | Dean, A. J., Bellgrove, M. A., Hall, T., Phan, W. M. J., Eyles, D. W., Kvaskoff, D., & McGrath, J. J. (2011). *Effects of Vitamin D Supplementation on Cognitive and Emotional Functioning in Young Adults – A Randomised Controlled Trial. PLoS ONE, 6(11), e25966.* |
| 73 | Diamond, T., Wong, Y. K., & Golombick, T. (2012). *Effect of oral cholecalciferol 2,000 versus 5,000 IU on serum vitamin D, PTH, bone and muscle strength in patients with vitamin D deficiency. Osteoporosis International, 24(3), 1101–1106.* |
| 74 | Close, G. L., Russell, J., Cobley, J. N., Owens, D. J., Wilson, G., Gregson, W., … Morton, J. P. (2013). Assessment of vitamin D concentration in non-supplemented professional athletes and healthy adults during the winter months in the UK: implications for skeletal muscle function. Journal of Sports Sciences, 31(4), 344–353. |

| | |
|---|---|
| 75 | Meekins, M. E., Oberhelman, S. S., Lee, B. R., Gardner, B. M., Cha, S. S., Singh, R. J., … Thacher, T. D. (2014). *Pharmacokinetics of daily versus monthly vitamin D3 supplementation in non-lactating women. European Journal of Clinical Nutrition, 68(5), 632–634.* |
| 76 | He, C.-S., Fraser, W. D., Tang, J., Brown, K., Renwick, S., Rudland-Thomas, J., … Gleeson, M. (2015). *The effect of 14 weeks of vitamin D3 supplementation on antimicrobial peptides and proteins in athletes. Journal of Sports Sciences, 34(1), 67–74.* |
| 77 | Nazarian, S., St. Peter, J. V., Boston, R. C., Jones, S. A., & Mariash, C. N. (2011). *Vitamin D3 supplementation improves insulin sensitivity in subjects with impaired fasting glucose. Translational Research, 158(5), 276–281.* |

### Supplementary Figures

**Figure S1.** Markov chain Monte Carlo simulation of the final model

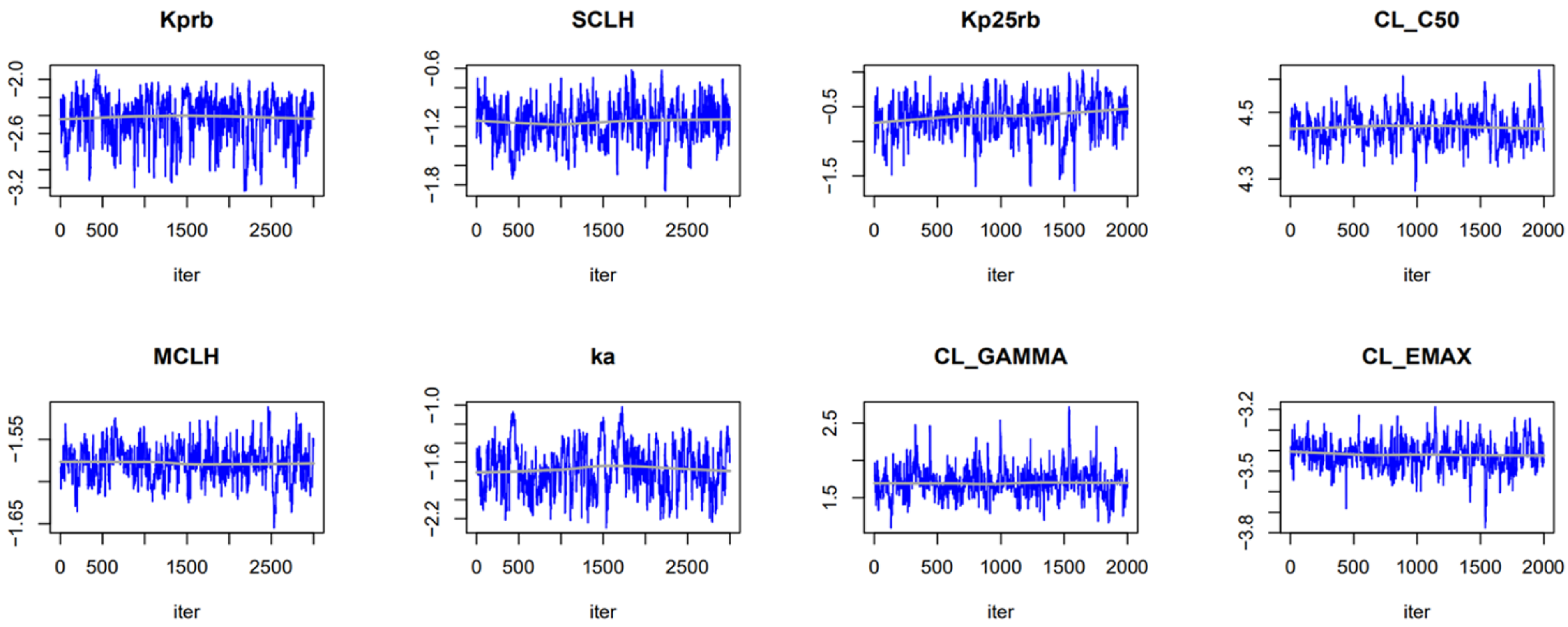

**Figure S2.** Visual predictive check of vitamin D after single dose and repeat dose.

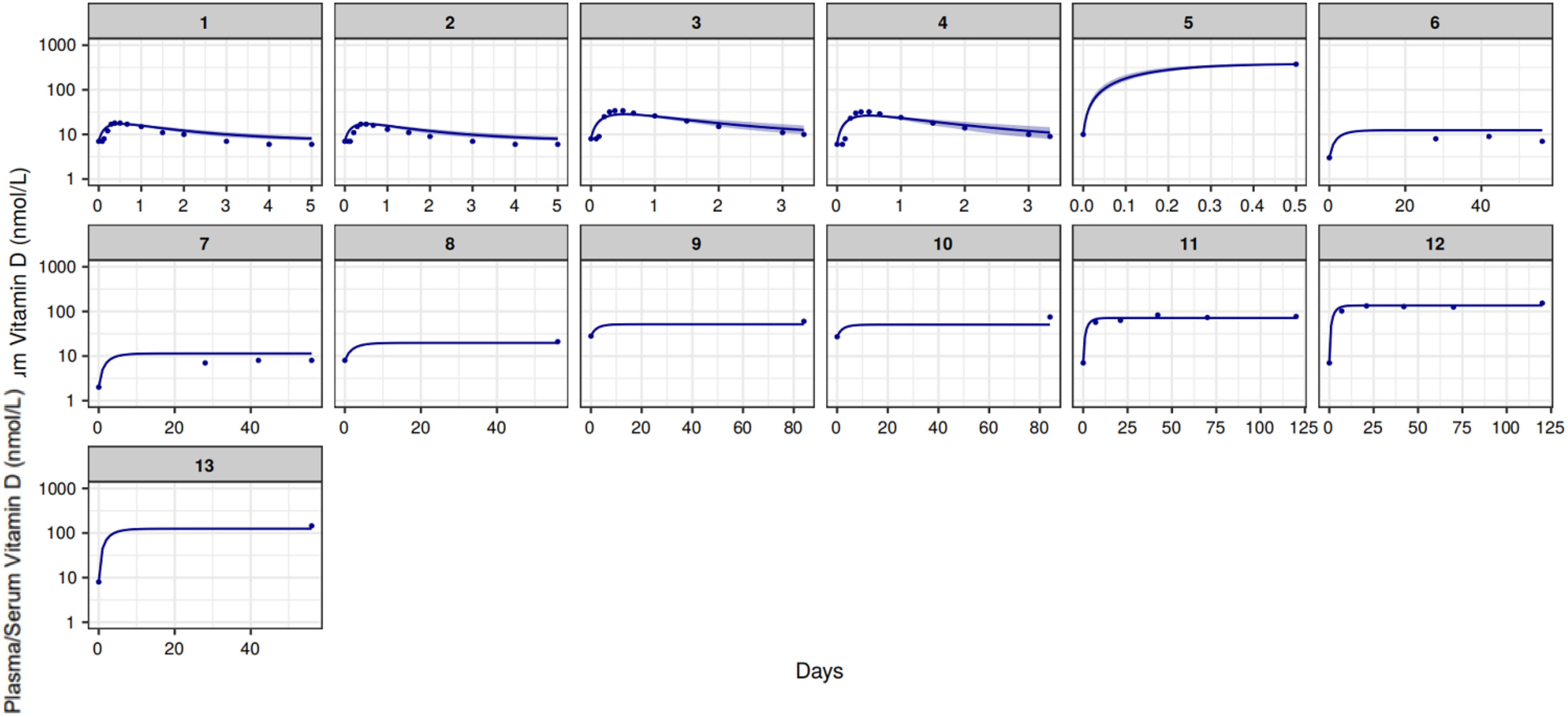

**Figure S3.** Visual predictive check of 25(OH)D after single dose

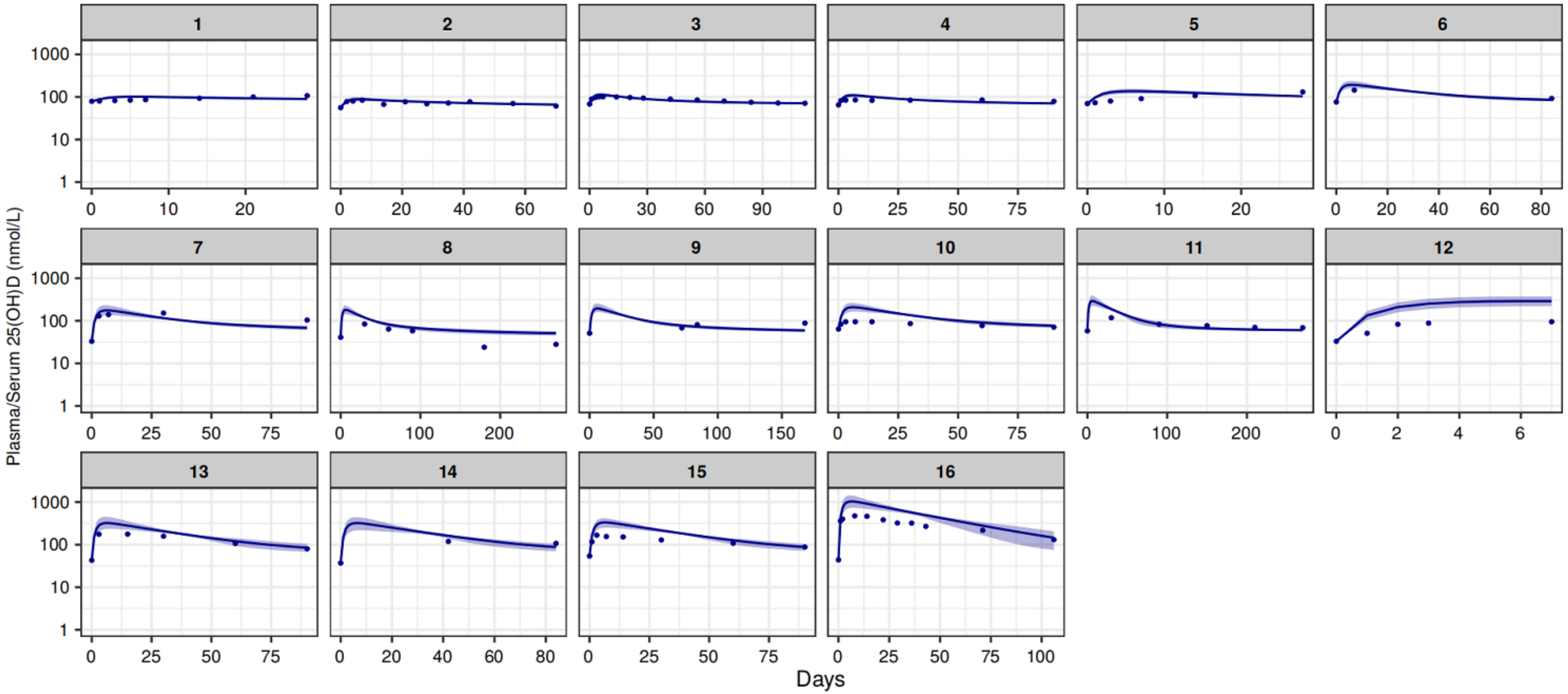

**Figure S4.** Visual predictive check of 25(OH)D after repeat dose of test set (all doses except 10μg and 100μg)

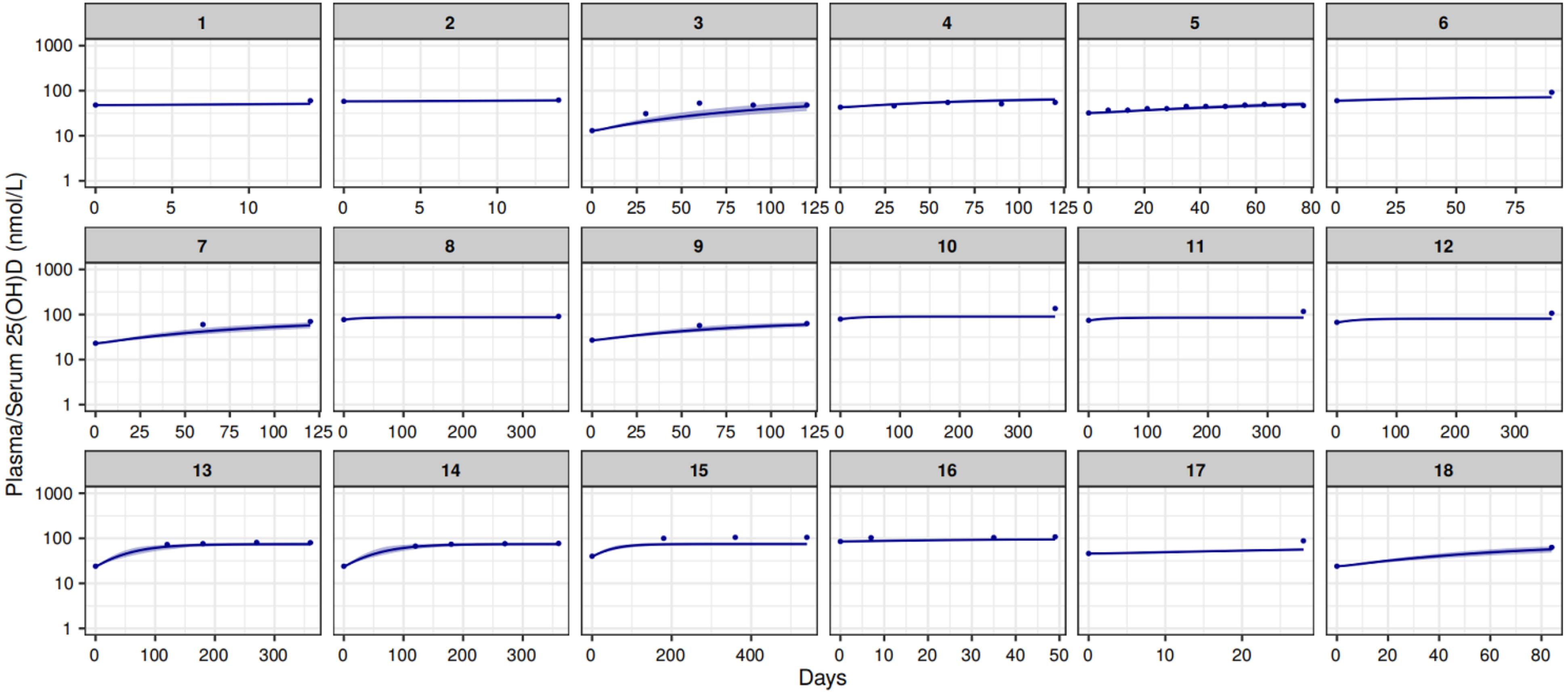

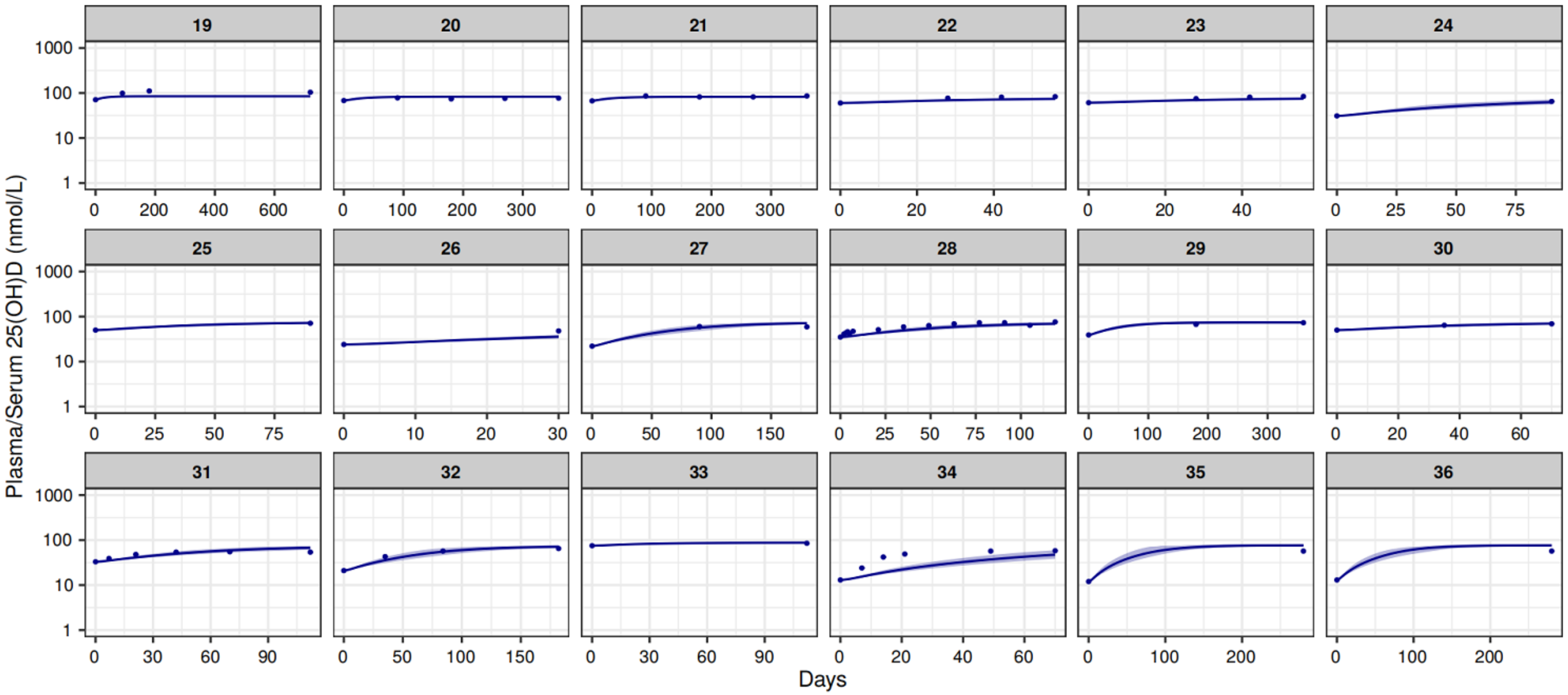

Plasma/Serum 25(OH)D (nmol/L)
Days

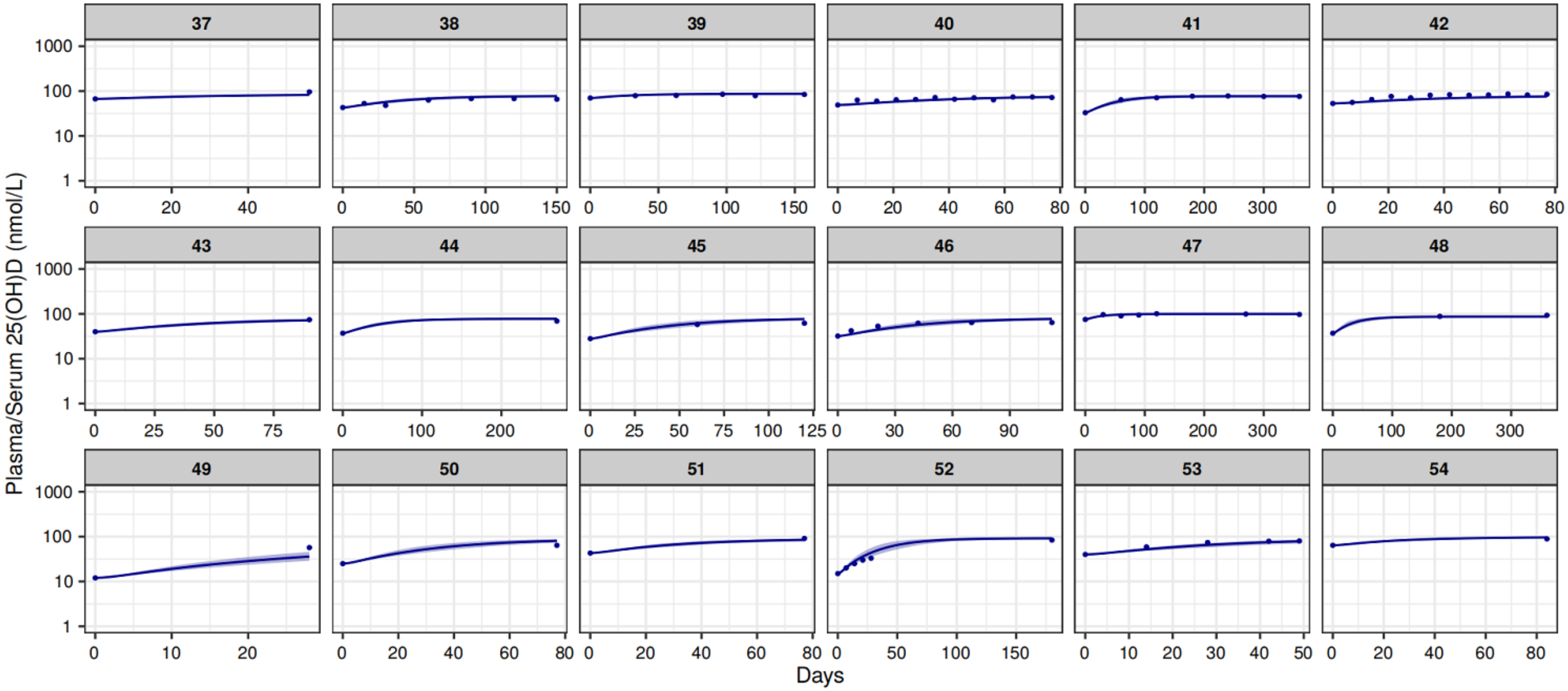

Plasma/Serum 25(OH)D (nmol/L)
Days

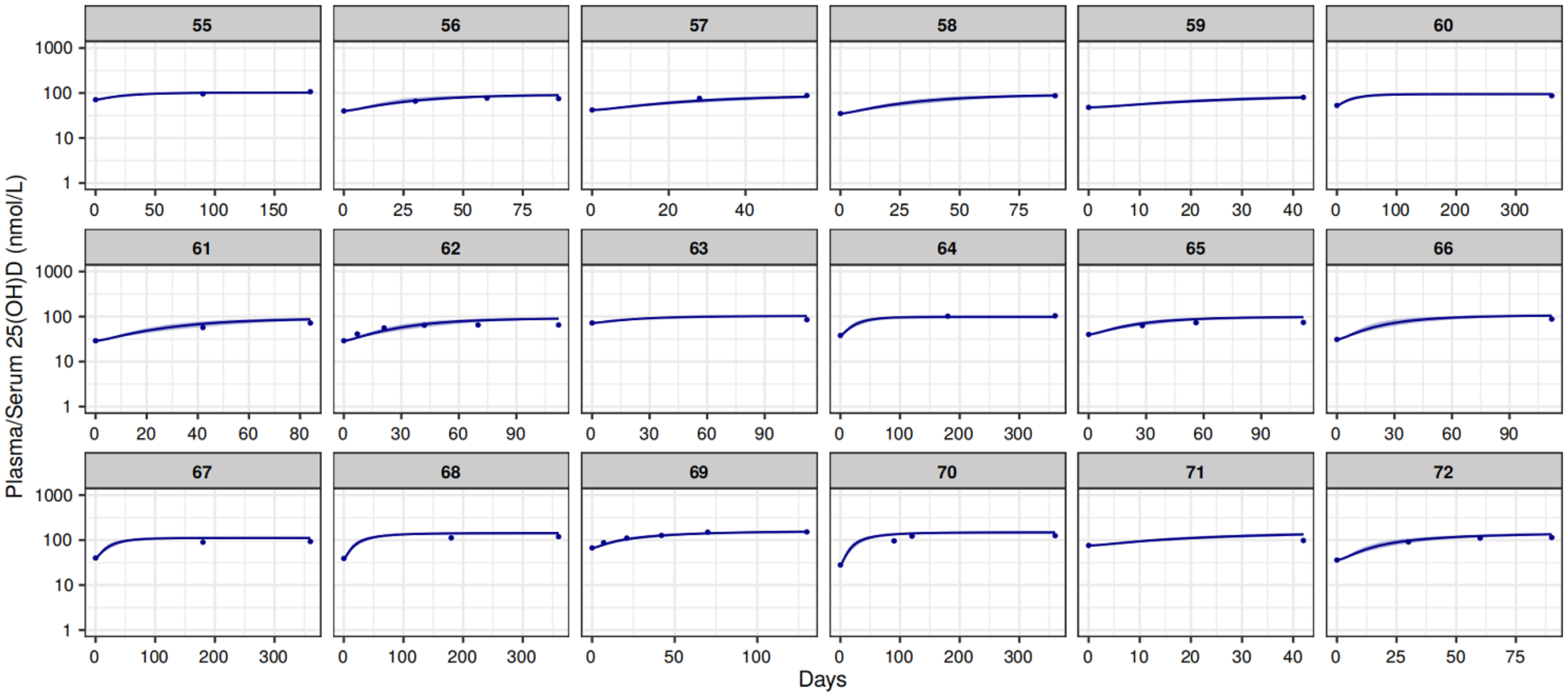

1000
100
10
1
Plasma/Serum 25(OH)D (nmol/L)
Days

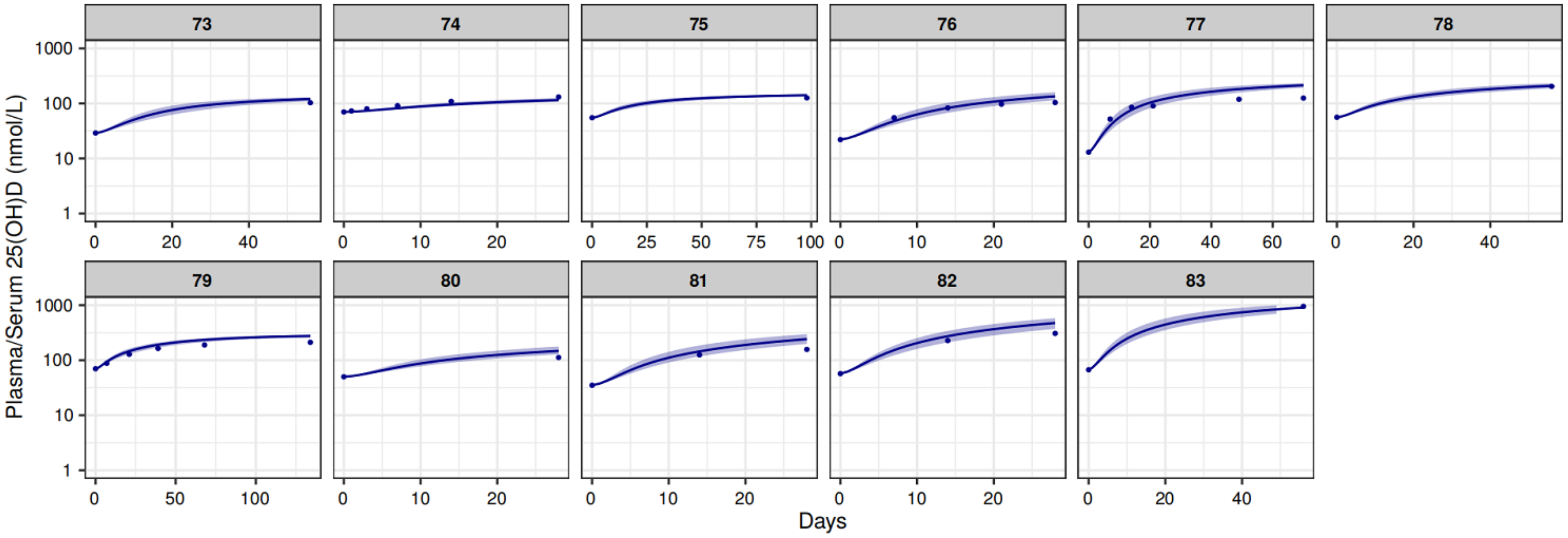

Plasma/Serum 25(OH)D (nmol/L)
1000
100
10
1
0
20
40
10
25
50
75
100
60
Days

**Figure S5.** Visual predictive check of 25(OH)D after 25(OH)D repeat dose

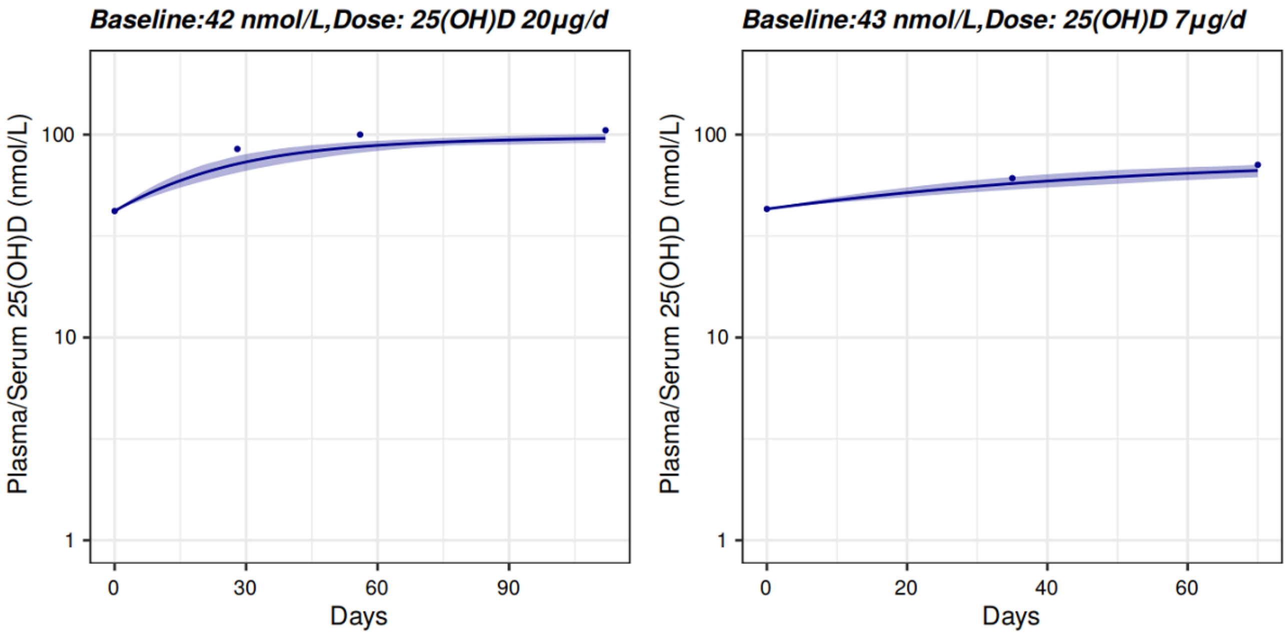

The observed data of the two cases derived from two references (Himmelstein, S., Clemens, T. L., Rubin, A., & Lindsay, R. (1990). Vitamin D supplementation in elderly nursing home residents increases 25(OH)D but not 1,25(OH)2D. The American Journal of Clinical Nutrition, 52(4), 701–706. Cherniack, E. P., Florez, H. J., Hollis, B. W., Roos, B. A., Troen, B. R., & Levis, S. (2011). *The Response of Elderly Veterans to Daily Vitamin D3 Supplementation of 2,000 IU: A Pilot Efficacy Study. Journal of the American Geriatrics Society, 59(2), 286–290.)*

**Figure S6.** Diagrams of each model structure.

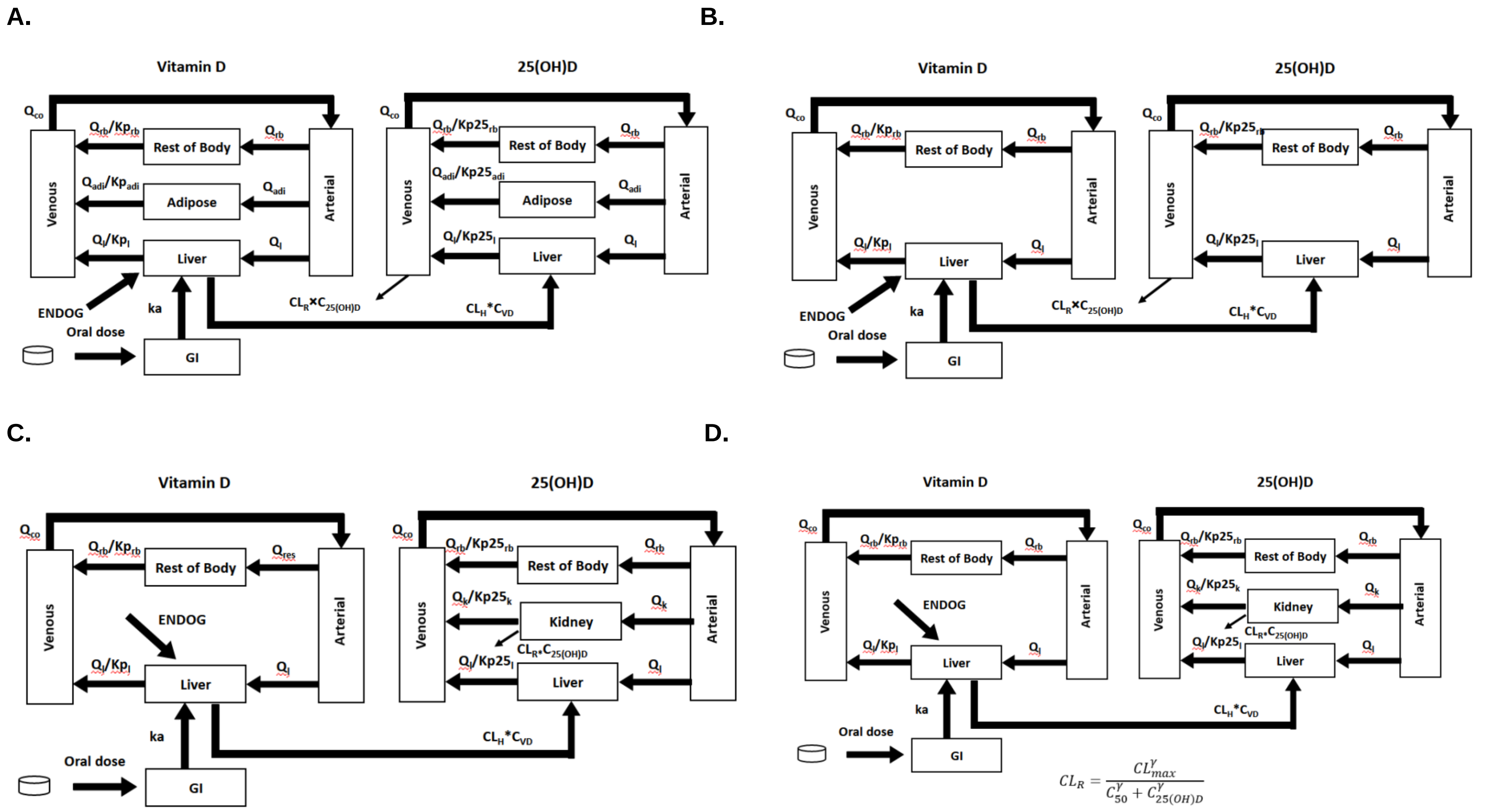

**E.**

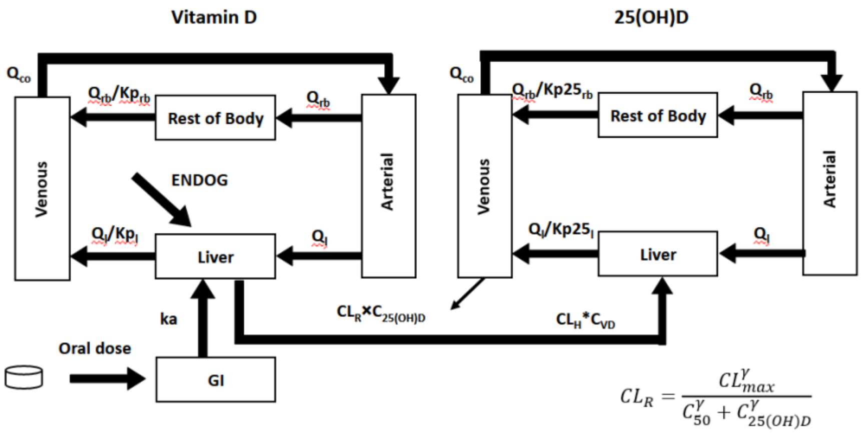

**F.**

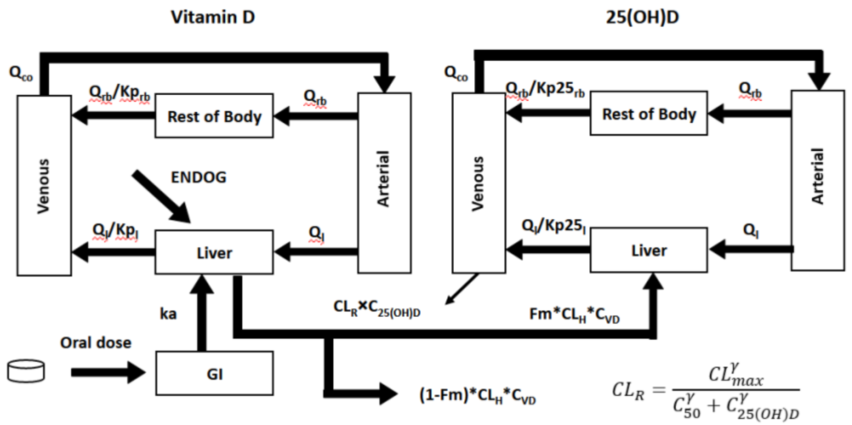

**G.**

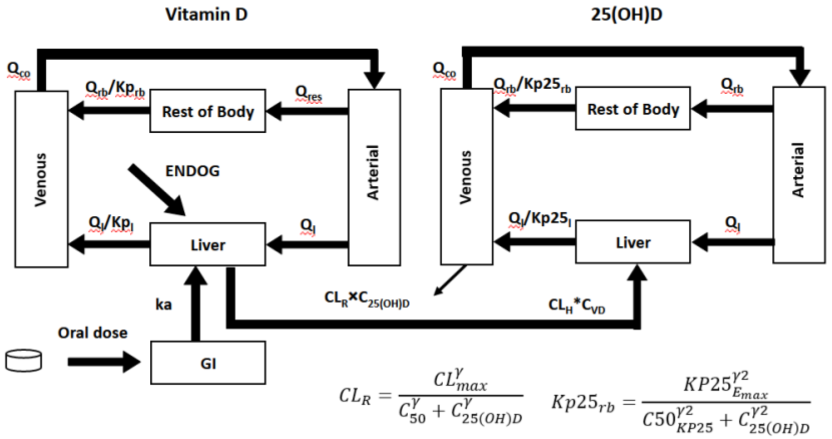

* Model structure for A) run001a, run001b, run001c, run002; B) run 003, run 004, run005, run006; C) run007; D) run008; E) run009 F) run009b G) run010.
